# Associations of drinking water per- and polyfluoroalkyl substances with cause-specific mortality in men with prostate cancer

**DOI:** 10.64898/2026.11.10.26367726

**Authors:** Hari S. Iyer, Megan R. Shanahan, Stefanie A. Joseph, Julianne Varga, Mike Z. He, Saum Ghodoussipour, Antoinette M. Stroup, Gerald Harris, Panos Georgopoulos, Robert J. Laumbach, Veronica M. Vieira, Jaime E. Hart, Emily S. Barrett, Elisa V. Bandera

## Abstract

**Background:** Over 3.5 million US men are living with prostate cancer (PCa). Certain per- and polyfluoroalkyl substances (PFAS) are carcinogenic and impair cardiometabolic health. Due to comorbidities and treatment, men with PCa may be uniquely susceptible to PFAS.

**Objectives:** To develop and validate a model for drinking water perfluorooctanoic acid (PFOA), perfluorooctane sulfonate (PFOS), and perfluorononanoic acid (PFNA), and study associations of each PFAS with all-cause, PCa, and cardiovascular disease (CVD) mortality in PCa survivors.

**Methods:** We conducted a cohort study of 51,289 men with PCa diagnosed from 2006 to 2017 and followed through 2024 from the New Jersey (NJ) cancer registry. PFAS monitoring data (n=26,161 results) were obtained from NJ government evaluations and Community Water Systems (CWS) compliance reports and were used to estimate PFAS concentrations using a two-phase XGBoost model with 67 predictors of PFAS contamination across 566 CWS. PFAS predictions between 2006-2024 were linked to participants’ address histories. Adjusted hazard ratios (aHR) and 95% confidence intervals (CI) were estimated from Cox models with sociodemographic, clinical and time-varying neighborhood covariates.

**Results:** There were 19,749 deaths, including 3,778 PCa deaths, and 4,972 CVD deaths over 466,843 person-years. Higher PFOA exposure was associated with all-cause mortality in certain quintiles (aHR_Q2vsQ1_: 1.07, 95% CI: 1.07-1.12; aHR_Q3vsQ1_: 1.06, 95% CI: 1.01-1.11) and with CVD mortality (aHR_Q5vsQ1_: 1.12, 95% CI: 1.01-1.24). Higher PFNA exposure was suggestively associated with all-cause mortality (aHR_≥3.8ng/L vs <2ng/L_: 1.22, 95% CI: 0.98-1.50) and PCa mortality (aHR_≥3.8ng/L vs <2ng/L_: 1.42, 95% CI: 0.96-2.10). Associations were strengthened when applying five-year lags. For example, higher PFOA (aHR_Q5vsQ1_: 1.15, 95% CI: 1.06-1.25), PFOS (aHR_Q5vsQ1_: 1.16, 95% CI: 1.07-1.25) and PFNA (aHR_≥3.8ng/Lvs <2ng/L_: 1.57, 95% CI: 1.06-2.31) were associated with higher all-cause mortality.

**Discussion:** Exposure to CWS-level PFAS was associated with higher mortality in PCa survivors. Replication is needed to confirm findings.

## Introduction

Prostate cancer (PCa) is a leading cause of cancer-related morbidity and mortality in US men, with 333,830 cases and 36,320 deaths each year^1^. There are 3.5 million PCa survivors in the US and although 10-year survival approaches 100%^2,3^, cardiovascular disease (CVD) is a major competing risk of death^4,5^, exceeding PCa mortality in men diagnosed with low-risk disease^6,7^. Men with PCa experience CVD risks from comorbidities, their advanced age, and treatment-related toxicities^6,8–10^. Despite this, clinical assessment of CVD risk factors is inconsistent, with roughly half of PCa survivors screened and managed for these conditions^11^. Improving CVD risk stratification, prevention and care in men with PCa has great potential for improving survivorship outcomes^12,13^.

Many environmental exposures are recognized as CVD risk factors^14,15^, but have only recently been incorporated into clinical risk prediction scores and other risk stratification tools in cancer survivors^4,14,16^. Drinking water contaminants, such as per- and polyfluoroalkyl substances (PFAS), are associated with higher lipid levels, impaired immune response, and epigenetic alterations in experimental and observational epidemiologic studies^17–22^. PFAS are a class of chemicals used in industrial manufacturing, consumer products, and firefighting foams which have received scrutiny due to their widespread occurrence and persistence in the environment and human bodies^23–25^. Perfluorooctanoic acid (PFOA) has been designated as carcinogenic to humans and perfluorooctanesulfonic acid (PFOS) as possibly carcinogenic to humans by the International Agency for Cancer Research (IARC) based on compelling mechanistic and animal experimental evidence, and limited human epidemiologic evidence for risks of renal cell carcinoma and testicular cancer associated with PFOA^26^. Results from epidemiologic studies of PFAS exposure and PCa risk have been mixed, possibly due to differences in exposure assessment and timing with respect to outcomes across studies^27–33^. Among adults in the general population, exposure to PFAS has been associated with higher lipid levels, but less consistently with CVD events^34–40^. Due to their higher burden of CVD risk factors than the general population^8^, PCa survivors could be particularly susceptible to CVD-related harms of PFAS. While evidence is accumulating for potential risks of PCa associated with PFAS exposure, there has been far less evidence regarding how PFAS may influence outcomes in PCa survivors.

New Jersey (NJ) was among the first states to conduct statewide surveillance and monitoring studies^41^, and to set Maximum Contaminant Levels for safe drinking water levels of perfluorooctanoic acid (PFOA), perfluorooctane sulfonic acid (PFOS), and perfluorononanoic acid (PFNA) between 2018 and 2020^42^. Although these regulations have led to reductions in drinking water PFAS concentrations^43^, because of the long half-lives of PFAS in human serum^44^, PCa survivors could still experience adverse health effects from earlier and ongoing exposure. We developed a geospatial exposure assessment approach to historically reconstruct PFOA, PFOS and PFNA concentrations in NJ drinking water. We studied associations of these modeled PFAS exposures with all-cause, PCa and CVD mortality, hypothesizing that men with higher drinking water PFAS exposure would experience higher mortality, particularly CVD mortality.

## Methods

### Study population and design

We conducted a cohort study using data from men with PCa from the NJ State Cancer Registry (NJSCR). The cohort included men diagnosed between January 1, 2006, and December 31, 2017, and followed until March 31, 2024. As part of the Surveillance, Epidemiology and End Results program, NJSCR follows systematic protocols to ensure data quality and completeness^45^. The registry captures demographic characteristics, tumor characteristics, first treatment, and date of diagnosis^46^.

The NJSCR collected residential histories through a linkage with the LexisNexis commercial database^47–49^. In brief, LexisNexis collects address information from real estate, mortgage, motor vehicle registration, driver license and other sources to enable greater capture of all addresses that an individual has lived in from 1980 onwards. Linkages were performed using a probabilistic matching algorithm Overall matching rates were nearly 98% for colorectal cancer^48^, and we observed 93% of addresses obtained through LexisNexis in this study of men with PCa. Prior to releasing the histories, officials from the cancer registry cleaned and processed addresses by removing duplicates and organizing by start and end dates, with the most recent address chosen in the case of overlapping periods. When addresses were missing over time intervals, we assumed the participant stayed at the same address^48^. Addresses were geomasked within 250m to protect patient privacy, following previously described procedures^50^. Specific details, including summaries of geocoding quality, number of addresses per study participant, and median duration of time at each address, are provided in the **Supplementary Methods**. Further details about the LexisNexis linkages using cancer registry data are available elsewhere^51,52^.

Of 131,091 individuals included in the NJSCR, we excluded men with no continuous NJ residence (n=30,421), those with diagnosis prior to 2006 (n=42,839), those diagnosed at autopsy (n=814), those for whom PFAS could not be linked (n=5,702), and those whose address history was incomplete (n=26) for an analytic sample of 51,289 (**Supplementary Figure 1**). The institutional review boards of the Dana-Farber Cancer Institute and Rutgers University, The State University, approved this study and determined that, because existing data sources were used, no written consent was required for participation in the study. Access to cancer registry data additionally required state institutional review board approval from NJ.

### Drinking water PFAS exposure assessment

An overview of the modeling strategy is presented here, with further details in **Supplementary Methods**. All predictor data, publicly available NJ PFAS monitoring data, and analysis code are publicly available through a GitHub repository^53^.

We predicted three-month average drinking water PFAS concentrations across 566 Community Water Systems (CWS) between January 2006 and December 2025. This start date was used because that was when the New Jersey Department of Environmental Protection first became aware of potential PFAS contamination in drinking water and began monitoring and surveillance^41^. We used the US Environmental Protection Agency’s modeled CWS boundaries to define water system areas^54^. This spatial scale was selected as the smallest geographic scale at which NJ drinking water monitoring data are reported.

PFAS monitoring data were obtained from the NJ Department of Environmental Protection’s historical sampling from 52 unique water systems between 2006 and 2010^41,55^, as well as the NJ Department of Environmental Protection’s “Drinking Water Watch” database, where public CWS are required to report on PFAS levels from routine testing as part of compliance with state regulations^56^. In addition to databases of NJ statewide monitoring and evaluation, we also obtained data from the Unregulated Contaminant Monitoring Rule (UCMR) conducted between 2013-2015 (UCMR3), and 2023-2026 (UCMR5) covering 263 unique water systems. Both UCMR3 and UCMR5 required all CWS and non-transient non-community water systems serving >10,000 people to report. For UCMR3, a nationally representative sample of 800 systems serving <3,300 or 3,300-<10,000 individuals was also included^57^. In UCMR5, all systems serving 3,300-<10,000 individuals were required to monitor, and a nationally representative sample of 800 water systems was selected to represent those serving <3,300 individuals^58^.

After exclusions of PFOA, PFOS and PFNA results from unfinished samples or those with data quality issues, we obtained 8,800 PFOA, 8,674 PFOS, and 8,687 PFNA results from these sources. The major assays used were EPA Methods 537 and 533. Limits of Detection for each source, along with PFAS assessment method, are provided in **Supplementary Table 1**.

We selected predictors based on literature describing sources of drinking water PFAS contamination and methodological papers aiming to predict PFAS concentrations in groundwater systems^59–61^. We identified military bases and training sites, airports, locations of businesses that have been associated with PFAS contamination, firefighting stations, neighborhood socioeconomic status (nSES), and outdoor air pollution (fine particulate matter, nitrogen dioxide) as a proxy for industrial contamination in the state of NJ. In addition, we obtained hydrological measures of clay, well depth, depth to water, estimated nitrogen loading, and groundwater discharge from gridded databases prepared by Tokranov et al. as part of a modeling study to predict US PFAS groundwater contamination^59^. Time-varying nSES and outdoor air pollution were updated (decennial census 2000 data were used for predictions between 2006 and 2009, and 2010 decennial census and American Community Survey data were used for 2010 onwards; air pollution was updated annually with a linear interpolation for years after 2016, the last year data were available). For point locations, we calculated proximity-based metrics for the number of locations (5, 10, 15) within pre-specified distances (1km, 5km, 10km) using ArcGIS (Redlands, CA). Our final predictor set included 67 measures (**Supplementary Table 2**). Specific business locations and codes are provided in **Supplementary Table 3**.

We then fit two-phase XGBoost models using 10-fold cross-validation with a 70%/30% training/test split in our primary analysis to ensure sufficient test data for drinking water contaminant prediction^62^. In the first phase, we predicted detectable vs non-detectable concentrations using a logistic regression classifier. In the second phase, we predicted average concentrations using a linear regression classifier. Hyperparameters were tuned using grid searches within the xgboost package in R, and final sets are provided in **Supplementary Table 4**.

We evaluated performance of the XGBoost models using Area Under the Curve (AUC) for the detect vs non-detect results, and the Root Mean Squared Error (RMSE), and R^2^ for concentrations (**Supplementary Table 5**). Training AUC for detect vs non-detect PFOA (0.94), PFOS (0.94), and PFNA (0.95) were excellent, and declined slightly in test samples (PFOA: 0.91, PFOS: 0.90, PFNA: 0.92). For PFAS concentrations, R^2^ were generally acceptable (PFOA: 0.75, PFOS: 0.64, PFNA: 0.88). These R^2^ declined in test samples (PFOA: 0.68, PFOS: 0.53, PFNA: 0.75) but remained acceptable and within the range of reported R^2^ from other drinking water exposure models^62^.

As a source of external validation, we compared modeled drinking water PFAS predictions and tap water collection from a pilot study of 71 NJ men with PCa^63^ within the same 3-month period. We found moderate correlations for PFOA (r=0.44, p<.001) and PFOS (r=0.33, p <.01) but weaker correlations for PFNA (r=0.03), which may have been due to the strong skew in PFNA drinking water predicted values.

Correlations between predictors and modeled PFAS were not indicative of collinearity (**Supplementary Figure 2**). Feature importance for models revealed that PM_2.5_ and NO_2_ were strong predictors for all models, along with certain measures of proximity to businesses known to produce PFAS and military bases (**Supplementary Figure 3, Supplementary Figure 4, Supplementary Figure 5**). Final exposure values were assigned to participants based on the CWS containing their residence in a given quarter. If participants changed addresses during a quarter, we applied a time-weighted average based on the number of days at each address.

We conducted sensitivity analyses using different training/test splits based on time period and CWS population density. These splits resulted in poor performance across validation metrics (**Supplementary Table 4, Supplementary Table 5**).

### Outcomes

Mortality was assessed from linkages between NJ state and national death records. ICD-09 and ICD-10 codes were used to categorize PCa-specific and CVD-specific causes of death^46^. Surveillance, Epidemiology and End Results algorithms were applied to address potential cancer-specific outcome misclassification per standard practice^64^.

### Covariates

We selected covariates using directed acyclic graphs^65^, specifying sociodemographic, clinical, and geospatial characteristics that would be correlated with residential drinking water PFAS and mortality. Our mortality models were adjusted for age at diagnosis (categorical: ≤50, 51-60, 61-70, 71-80, ≥80 years), diagnosis year (categorical: 2006-2009, 2010-2013, 2014-2017), race and ethnicity (categorical: non-Hispanic White, non-Hispanic Black, Hispanic, Other (non-Hispanic American Indian, Alaska Native, Pacific Islander)), marital status (binary), insurance (categorical: uninsured, private, Medicaid, Medicare, Other government, Unknown), stage at diagnosis (categorical: Localized, Regional, Distant), receipt of surgery (binary), receipt of radiotherapy (binary), and prostate-specific antigen (PSA) at diagnosis (categorical: <10, 10-<20, ≥20 ng/mL). In addition, we adjusted for time-varying tract-level neighborhood socioeconomic status (quintiles)^66^, residential address-level outdoor fine particulate matter (continuous), outdoor nitrogen dioxide (continuous), and residential address-level Normalized Difference Vegetation Index (NDVI) within 270m as a measure of greenspace^67^.

### Statistical analysis

We studied correlations of drinking water PFAS exposures and covariates using chi-squared tests for categorical variables, two-sample independent t-tests for means, and non-parametric Wilcoxon rank sum tests for skewed continuous measures. We fit Kaplan-Meier curves for associations of drinking water PFOA and PFOS quintiles and overall survival and used log-rank tests to assess significant differences in survival curves. Due to high level of skew, we categorized PFNA as <2 ng/L, 2-<3.8 ng/L, and ≥3.8 ng/L using the median of the distribution of continuous PFNA values. For multivariable analyses, we fit time-dependent Cox proportional hazards models based on 3-month intervals aligning with the resolution of our drinking water PFAS models, with time-varying PFAS exposure and nSES, greenspace, and outdoor air pollution covariates.

We performed sensitivity analyses to assess whether five-year lagged exposures had stronger associations with the outcomes, based on the assumption that there would be a delayed effect of PFAS exposure on our outcomes of interest. We restricted the population to 28,840 individuals diagnosed from January 2011 onwards for whom a minimum of five years of PFAS exposure data were available. We then fit our main Cox models with five-year lags also applied to time-varying covariates (PM_2.5_, NO_2_, nSES, and NDVI).

In order to assess whether specific PFAS cutpoints could confer elevated risks and to reduce influence of outliers, PFAS were parametrized as quintiles with a test for ordinal trend, along with a continuous measure per interquartile range increase. We further assessed non-linearity using restricted cubic splines^68^, but did not find clear evidence of non-linearity based on likelihood ratio tests. Primary analyses were conducted separately for PFOA, PFOS and PFNA. We used quantile g-computation to estimate the joint impacts of all three PFAS as a mixture, estimating the effect of a one-quintile increase across all PFAS^69^. We additionally assessed effect modification of the association per one IQR increase in PFOA, PFOS and PFNA with each mortality endpoint by age, race, and stage using multiplicative interaction terms and likelihood ratio tests.

For PCa and CVD mortality, we used cause-specific models to account for competing risks, assuming non-informative censoring through non-PCa or CVD death^70^. As sensitivity analysis, we used inverse probability of censoring (IPCW) weights to model associations of drinking water PFAS and PCa and CVD death, assuming no deaths from the competing cause^71^.

Several covariates had substantial missingness (as high as 22-23% for stage and PSA value at diagnosis). For geospatial time-varying variables, we first used linear interpolation to predict values outside of the range of available data at the participant’s address from 2006 to 2025. Next, for individual sociodemographic and clinical factors, we performed multiple imputation with chained equations using the mice package in R. We used predictive mean matching algorithms for continuous data, logistic regression for binary data, and multinomial logistic regression for categorical data, using covariate data only for imputation models. We used 5 imputed datasets and Rubin’s Rules to pool and correct standard errors^72^.

All tests were two-sided with α=0.05. Analyses were conducted using R version 4.5.2, with spatial analyses for inverse distance processing using ArcGIS Pro.

## Results

### Participant characteristics

Characteristics of the 51,289 men with PCa diagnosed from 2006-2017 who met eligibility criteria are presented in **Table 1**, with summary statistics for PFOA (median: 1.65, IQR [1.12-4.87 ng/L]), PFOS (median: 1.13, IQR [1.02-1.86 ng/L]) and PFNA (median: 1.01, IQR [1.00-1.02]) drinking water concentrations presented in **Table 2**. Mean age at diagnosis was 67.1 years (SD=9.4), 70% were non-Hispanic White (NHW), 16% were non-Hispanic Black (NHB), 4% were non-Hispanic Asian, Pacific Islander or Alaska Native, and 10% were Hispanic. Of those with complete PCa stage information, 87% were diagnosed with localized disease, 8% with regional disease, and 5% with distant disease. Men residing in neighborhoods served by CWS in the highest compared to lowest quintiles of PFOA were more likely to have been diagnosed earlier (2006-2010: 55% vs 31%), be NHB (21% vs 12%) or Hispanic (14% vs 5%), and slightly more likely reside in the highest (most advantaged) quintile of nSES (26% vs 20%). These associations between sociodemographic factors and drinking water PFOS were largely similar (**Supplementary Table 6**). However, for PFNA (**Supplementary Table 7**), there was no association between CWS PFNA levels and year of diagnosis, and less clear associations with respect to race and ethnicity. In addition, those served by CWS with the highest compared to lowest PFNA levels were more likely to reside in the lowest (most deprived) quintile of nSES (49% vs 15%).

**Table 1.** Characteristics of New Jersey Men with Prostate Cancer Stratified by Quintile of PFOA.

|  | Quintiles of PFOA |  |  |  |  |  |
|---|---|---|---|---|---|---|
| Characteristic (n, %) | Quintile 1 | Quintile 2 | Quintile 3 | Quintile 4 | Quintile 5 | Overall |
| **N** | **12024** | **11291** | **8258** | **7940** | **11776** | **51289** |
| **PFOA, ng/L (median [IQR])** | 1.04 [1.02, 1.07] | 1.24 [1.16, 1.37] | 1.97 [1.65, 2.31] | 3.96 [3.26, 4.55] | 8.32 [6.27, 13.83] | 1.65 [1.12, 4.87] |
| **Age at diagnosis, years (mean, SD)** | 67.13 (9.33) | 66.91 (9.39) | 67.24 (9.43) | 67.11 (9.34) | 67.33 (9.46) | 67.14 (9.39) |
| **Age at diagnosis, years** |  |  |  |  |  |  |
| <50 | 395 (3%) | 408 (4%) | 290 (4%) | 256 (3%) | 394 (3%) | 1743 (3.4%) |
| 50-60 | 2533 (21%) | 2498 (22%) | 1726 (21%) | 1724 (22%) | 2522 (21%) | 11003 (21.5%) |
| 60-70 | 4792 (40%) | 4411 (39%) | 3248 (39%) | 3159 (40%) | 4519 (38%) | 20129 (39.2%) |
| 70-80 | 3338 (28%) | 3107 (28%) | 2267 (27%) | 2138 (27%) | 3300 (28%) | 14150 (27.6%) |
| ≥80 | 966 (8%) | 867 (8%) | 727 (9%) | 663 (8%) | 1041 (9%) | 4264 (8.3%) |
| **Year of diagnosis** |  |  |  |  |  |  |
| 2006-2010 | 3747 (31%) | 3510 (31%) | 2940 (36%) | 2038 (26%) | 6467 (55%) | 18702 (36.5%) |
| 2010-2014 | 5169 (43%) | 4385 (39%) | 2026 (25%) | 3123 (39%) | 1865 (16%) | 16568 (32.3%) |
| 2014 -2017 | 3108 (26%) | 3396 (30%) | 3292 (40%) | 2779 (35%) | 3444 (29%) | 16019 (31.2%) |
| **Race/Ethnicity** |  |  |  |  |  |  |
| Non-Hispanic White | 9579 (80%) | 8120 (72%) | 5764 (70%) | 5194 (65%) | 7246 (62%) | 35903 (70.0%) |
| Non-Hispanic Black | 1450 (12%) | 1858 (16%) | 1270 (15%) | 1375 (17%) | 2445 (21%) | 8398 (16.4%) |
| Non-Hispanic Asian American/Pacific Islander | 396 (3%) | 349 (3%) | 360 (4%) | 390 (5%) | 482 (4%) | 1977 (3.9%) |
| Hispanic | 599 (5%) | 964 (9%) | 864 (10%) | 981 (12%) | 1603 (14%) | 5011 (9.8%) |
| **Marital Status** |  |  |  |  |  |  |
| No | 2238 (19%) | 2364 (21%) | 1670 (20%) | 1679 (21%) | 2518 (21%) | 10469 (20.4%) |
| Yes | 7861 (65%) | 6960 (62%) | 5057 (61%) | 4834 (61%) | 7400 (63%) | 32112 (62.6%) |
| Missing | 1925 (16%) | 1967 (17%) | 1531 (19%) | 1427 (18%) | 1858 (16%) | 8708 (17.0%) |
| **Insurance** |  |  |  |  |  |  |
| Uninsured | 768 (6%) | 824 (7%) | 549 (7%) | 484 (6%) | 832 (7%) | 3457 (6.7%) |
| Private | 3653 (30%) | 3238 (29%) | 2245 (27%) | 2296 (29%) | 3165 (27%) | 14597 (28.5%) |
| Medicaid | 144 (1%) | 177 (2%) | 137 (2%) | 129 (2%) | 196 (2%) | 783 (1.5%) |
| Medicare | 4929 (41%) | 4288 (38%) | 3056 (37%) | 2804 (35%) | 4237 (36%) | 19314 (37.7%) |
| Other government | 35 (0%) | 32 (0%) | 16 (0%) | 12 (0%) | 41 (0%) | 136 (0.3%) |
| Unknown | 2495 (21%) | 2732 (24%) | 2255 (27%) | 2215 (28%) | 3305 (28%) | 13002 (25.4%) |
| **Gleason (combined)** |  |  |  |  |  |  |
| <7 | 6305 (52%) | 6206 (55%) | 4694 (57%) | 4486 (56%) | 6460 (55%) | 28151 (54.9%) |
| ≥7 | 4647 (39%) | 4112 (36%) | 2887 (35%) | 2788 (35%) | 4449 (38%) | 18883 (36.8%) |
| Unknown | 1072 (9%) | 973 (9%) | 677 (8%) | 666 (8%) | 867 (7%) | 4255 (8.3%) |
| **Stage** |  |  |  |  |  |  |
| Localized | 8698 (72%) | 8247 (73%) | 5352 (65%) | 5202 (66%) | 7424 (63%) | 34923 (68.1%) |
| Regional | 905 (8%) | 772 (7%) | 544 (7%) | 468 (6%) | 653 (6%) | 3342 (6.5%) |
| Distant | 479 (4%) | 430 (4%) | 280 (3%) | 260 (3%) | 334 (3%) | 1783 (3.5%) |
| Unknown | 1942 (16%) | 1842 (16%) | 2082 (25%) | 2010 (25%) | 3365 (29%) | 11241 (21.9%) |
| **Surgery** |  |  |  |  |  |  |
| No | 7719 (64%) | 7536 (67%) | 5399 (65%) | 5254 (65%) | 7807 (66%) | 33715 (65.7%) |
| Yes | 4124 (34%) | 3593 (32%) | 2709 (33%) | 2489 (31%) | 3698 (31%) | 16613 (32.4%) |
| Unknown | 181 (2%) | 162 (1%) | 150 (2%) | 197 (2%) | 271 (2%) | 961 (1.9%) |
| **Hormone treatment** |  |  |  |  |  |  |
| No | 8803 (73%) | 8146 (72%) | 5852 (71%) | 5605 (71%) | 7822 (66%) | 36228 (70.6%) |
| Yes | 2914 (24%) | 2921 (26%) | 2229 (27%) | 2104 (26%) | 3573 (30%) | 13741 (26.8%) |
| Unknown | 307 (3%) | 224 (2%) | 177 (2%) | 231 (3%) | 381 (3%) | 1320 (2.6%) |
| **Radiation** |  |  |  |  |  |  |
| No | 6961 (58%) | 6255 (55%) | 4752 (58%) | 4507 (57%) | 6603 (56%) | 29078 (56.7%) |
| Yes | 4918 (41%) | 4891 (43%) | 3418 (41%) | 3319 (42%) | 5077 (43%) | 21623 (42.2%) |
| Unknown | 145 (1%) | 145 (1%) | 88 (1%) | 114 (1%) | 96 (1%) | 588 (1.1%) |
| **PSA Category, ng/mL** |  |  |  |  |  |  |
| <10 | 7775 (65%) | 6994 (62%) | 4802 (58%) | 4645 (59%) | 6987 (59%) | 31203 (60.8%) |
| 10-20 | 1198 (10%) | 1108 (10%) | 862 (10%) | 793 (10%) | 1246 (11%) | 5207 (10.2%) |
| ≥20 | 636 (5%) | 655 (6%) | 478 (6%) | 490 (6%) | 721 (6%) | 2980 (5.8%) |
| Unknown | 2415 (20%) | 2534 (22%) | 2116 (26%) | 2012 (26%) | 2822 (24%) | 11899 (23.2%) |
| **Neighborhood SES (Quintile)** |  |  |  |  |  |  |
| Quintile 1 (Most deprived) | 2551 (21%) | 2013 (18%) | 1168 (14%) | 886 (11%) | 1624 (14%) | 8242 (16.1%) |
| Quintile 2 | 2309 (19%) | 2309 (20%) | 1484 (18%) | 1220 (15%) | 2202 (19%) | 9524 (18.6%) |
| Quintile 3 | 2290 (19%) | 2416 (21%) | 1691 (20%) | 1349 (17%) | 2489 (21%) | 10235 (20.0%) |
| Quintile 4 | 2460 (20%) | 2297 (20%) | 2018 (24%) | 2190 (28%) | 2457 (21%) | 11422 (22.3%) |
| Quintile 5 (Least deprived) | 2414 (20%) | 2256 (20%) | 1897 (23%) | 2295 (29%) | 3004 (26%) | 11866 (23.1%) |
| **County-level PSA, % (mean (SD))** | 0.57 (0.04) | 0.56 (0.05) | 0.57 (0.05) | 0.56 (0.05) | 0.58 (0.04) | 0.57 (0.04) |
| Unknown | 1398 (12%) | 1368 (12%) | 1795 (22%) | 1758 (22%) | 3097 (26%) | 9416 (18%) |
| **NO<sub>2</sub>, ppb (mean (SD))** | 24.66 (6.70) | 26.82 (5.76) | 28.37 (6.17) | 29.84 (7.15) | 31.94 (7.16) | 28.21 (7.12) |
| **PM<sub>2.5</sub>, µg/m<sup>3</sup> (mean (SD))** | 9.33 (1.58) | 9.20 (1.49) | 9.37 (1.72) | 9.43 (1.74) | 10.23 (2.06) | 9.53 (1.78) |
| **NDVI 270m (mean (SD))** | 0.36 (0.17) | 0.35 (0.17) | 0.34 (0.18) | 0.34 (0.17) | 0.31 (0.17) | 0.34 (0.17) |

Abbreviations: IQR=interquartile range; NDVI=Normalized Difference Vegetation Index; NO<sub>2</sub>=nitrogen dioxide; PFOA=perfluorooctanoic acid; PM<sub>2.5</sub>=fine particulate matter ≤2.5 microns in diameter; PSA=prostate-specific antigen; SD=standard deviation; SES=socioeconomic status

**Table 2.** Summary Statistics for drinking water PFOA, PFOS, and PFNA exposure in men with prostate cancer, New Jersey, 2006-2024.

| **Analyte** | **Minimum** | **p25** | **p50** | **p75** | **p90** | **Maximum** | **Mean** | **SD** |
|---|---|---|---|---|---|---|---|---|
| **PFOA, ng/L** | 1.00 | 1.12 | 1.65 | 4.87 | 9.08 | 52.03 | 3.94 | 5.15 |
| **PFOS, ng/L** | 0.99 | 1.02 | 1.13 | 1.86 | 4.79 | 25.72 | 2.36 | 3.42 |
| **PFNA, ng/L** | 0.98 | 1.00 | 1.01 | 1.02 | 1.07 | 34.65 | 1.09 | 0.74 |

### Maps of PFAS in NJ CWS

We mapped CWS-level PFAS predictions from our model in NJ for PFOA (**Supplementary Figure 6**), PFOS (**Supplementary Figure 7**), and PFNA (**Supplementary Figure 8**). Our maps showed declines in concentrations of these PFAS over time, but also evidence of potential stagnation in certain areas. For example, maps of predicted CWS PFOA levels showed persistently high concentrations in the northeastern part of NJ, although levels in other parts of the state declined. Similar patterns were observed for PFOS, but for PFNA, concentrations were high near Trenton and Paulsboro (in the southwestern part of NJ), an established contamination site^73^, and then declined.

### Kaplan-Meier survival

There were 19,749 all-cause deaths, 3,778 PCa deaths, and 4,972 CVD deaths over 466,843 person-years of follow-up. Kaplan-Meier curves presenting associations of PFOA, PFOS and PFNA with survival from all-cause mortality are presented in **Figure 1**. The median survival time for men was similar across quintiles of PFOA and PFOS. However, median survival time was shorter among those exposed to drinking water PFNA ≥3.8ng/L (12.92, 95% CI: 11.50, 14.34 years) vs <2 ng/L (14.59, 95% CI: 14.50, 14.75 years, log-rank *P* <.001).

**Figure 1.**
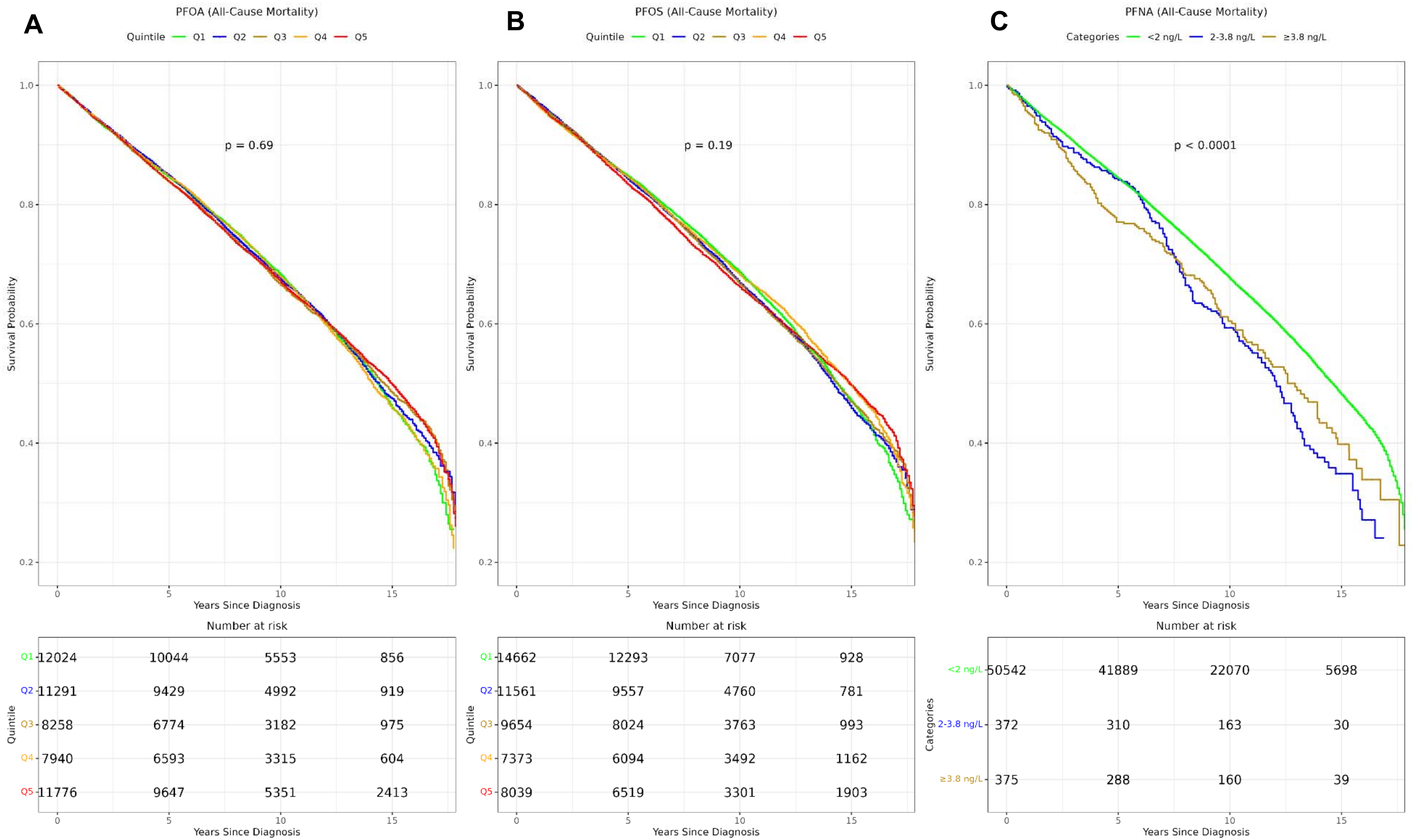
Kaplan-Meier plots of overall survival associated with PFOA (Panel A, Quintiles), PFOS (Panel B, Quintiles), and PFNA (Panel C, Categories: <2 ng/L, 2-<3.8 ng/L, ≥3.8 ng/L). Log-rank p-values are two-sided. PFOA=perfluorooctanoic acid, PFOS=perfluorooctane sulfonate, PFNA=perfluorononanoic acid.

### Multivariable analysis

After full covariate adjustment, there was no evidence of associations between continuous drinking water PFOA, PFOS or PFNA with all-cause mortality (**Table 3**). However, we observed higher mortality among certain quintiles of PFOA exposure (aHR_Q2vsQ1_: 1.07, 95% CI: 1.02, 1.12); aHR_Q3vsQ1_: 1.06, 95% CI: 1.01, 1.11; aHR_Q4vsQ1_: 1.05, 95% CI: 1.00, 1.10; aHR_Q5vsQ1_: 1.03, 95% CI: 0.98, 1.09). Compared to those exposed to <2 ng/L PFNA levels, those with PFNA ≥3.8 ng/L had higher mortality, though this result did not reach statistical significance (aHR: 1.42, 95% CI: 0.96, 2.10, *P*_trend_ = 0.06). In analyses using five-year lags, higher PFOA (aHR_Q5vsQ1_: 1.15, 95% CI: 1.06, 1.25, *P*_trend_ <.001), PFOS (aHR_Q5vsQ1_: 1.16, 95% CI: 1.07, 1.25, *P*_trend_ <.001), and PFNA ≥3.8 ng/L (aHR: 1.57, 95% CI: 1.06, 2.31, *P*_trend_ = 0.08) were associated with higher all-cause mortality, though analyses using quintiles revealed significantly lower mortality among those in quintile 3 for PFOA (aHR_Q3vsQ1_: 0.77, 95% CI: 0.71, 0.84) and PFOS (aHR_Q3vsQ1_: 0.84, 95% CI: 0.77, 0.91).

**Table 3.** Associations of drinking water PFOA, PFOS and PFNA with all-cause mortality in New Jersey men with prostate cancer.

|  |  | Quintiles of Exposure |  |  |  |  |  |
|---|---|---|---|---|---|---|---|
|  | **Continuous<sup>a</sup>** | **Quintile 1** | **Quintile 2** | **Quintile 3** | **Quintile 4** | **Quintile 5** | ***P*<sub>trend</sub>** |
|  | **aHR (95% CI)** | **aHR (95% CI)** | **aHR (95% CI)** | **aHR (95% CI)** | **aHR (95% CI)** | **aHR (95% CI)** |  |
| **Primary (n=51,289)** |  |  |  |  |  |  |  |
| PFOA |  |  |  |  |  |  |  |
| Cases/Person-years | 19,749/466,843 | 3,516/93,535 | 3,951/93,147 | 4,278/92,964 | 4,080/93,815 | 3,924/93,382 |  |
| Minimal<sup>b</sup> | 0.99 (0.97, 1.00) | Ref | 1.08 (1.03, 1.13) | 1.03 (0.98, 1.08) | 1.03 (0.99, 1.08) | 0.99 (0.95, 1.04) | 0.028 |
| Full<sup>c</sup> | 1.00 (0.99, 1.01) | Ref | 1.07 (1.02, 1.12) | 1.06 (1.01, 1.11) | 1.05 (1.00, 1.10) | 1.03 (0.98, 1.09) | 0.66 |
| PFOS |  |  |  |  |  |  |  |
| Cases/Person-years | 19,749/466,843 | 3,465/93,431 | 3,615/93,522 | 3,846/93,494 | 4,497/93,106 | 4,326/93,290 |  |
| Minimal<sup>b</sup> | 0.99 (0.98, 0.99) | Ref | 0.96 (0.92, 1.01) | 0.93 (0.88, 0.97) | 0.99 (0.95, 1.04) | 0.93 (0.89, 0.98) | 0.10 |
| Full<sup>c</sup> | 1.00 (0.99, 1.01) | Ref | 0.99 (0.94, 1.04) | 0.97 (0.92, 1.02) | 1.01 (0.96, 1.06) | 0.99 (0.94, 1.04) | 0.98 |
| PFNA<sup>d</sup> |  |  |  |  |  |  |  |
| Cases/Person-years | 19,749/466,843 | 19,589/463,111 | 73/1,933 | 87/1,800 | - | - |  |
| Minimal<sup>b</sup> | 1.00 (1.00, 1.00) | Ref | 1.17 (0.93, 1.47) | 1.41 (1.14, 1.75) | - | - | <.001 |
| Full<sup>c</sup> | 1.00 (1.00, 1.00) | Ref | 0.94 (0.75, 1.18) | 1.22 (0.98, 1.50) | - | - | 0.15 |
| **5-year lag (n=24,840)** |  |  |  |  |  |  |  |
| PFOA |  |  |  |  |  |  |  |
| Cases/Person-years | 7,157/190,028 | 1,371/38,338 | 1,489/38,310 | 1,005/38,029 | 1,605/37,648 | 1,687/37,702 |  |
| Minimal<sup>b</sup> | 1.02 (1.01, 1.04) | Ref | 1.08 (1.01, 1.18) | 0.77 (0.71, 0.84) | 1.05 (0.98, 1.13) | 1.08 (1.01, 1.16) | <.001 |
| Full<sup>c</sup> | 1.03 (1.01, 1.05) | Ref | 1.09 (1.01, 1.18) | 0.77 (0.71, 0.84) | 1.11 (1.03, 1.20) | 1.15 (1.06, 1.25) | <.001 |
| PFOS |  |  |  |  |  |  |  |
| Cases/Person-years | 7,157/190,028 | 1,297/38,559 | 1,417/38,430 | 1,086/38,041 | 1,540/37,973 | 1,817/37,024 |  |
| Minimal<sup>b</sup> | 1.01 (1.00, 1.01) | Ref | 1.04 (0.96, 1.12) | 0.84 (0.78, 0.91) | 1.01 (0.93, 1.08) | 1.10 (1.02, 1.19) | <.001 |
| Full<sup>c</sup> | 1.01 (1.00, 1.01) | Ref | 1.07 (0.99, 1.15) | 0.84 (0.77, 0.91) | 1.07 (0.99, 1.16) | 1.16 (1.07, 1.25) | <.001 |
| PFNA<sup>d</sup> |  |  |  |  |  |  |  |
| Cases/Person-years | 7,157/190,028 | 7,117/188,943 | 13/556 | 27/529 | - | - |  |
| Minimal<sup>b</sup> | 1.00 (1.00, 1.00) | Ref | 0.90 (0.52, 1.54) | 2.04 (1.40, 2.99) | - | - | .001 |
| Full<sup>c</sup> | 1.00 (1.00, 1.00) | Ref | 0.75 (0.44, 1.30) | 1.57 (1.06, 2.31) | - | - | 0.079 |

<sup>a</sup>Per interquartile range, <sup>b</sup>Adjusted for age, diagnosis year, <sup>c</sup>Adjusted for <sup>b</sup> and race/ethnicity, marital status, insurance status, stage, treatment (surgery and or radiation), PSA, nSES quintile, NO<sub>2</sub>, PM<sub>2.5</sub>, and NDVI (green space), <sup>d</sup>Due to skewness of PFNA measure, we used categorical cutoffs (<2 ng/L, 2-<3.8 ng/L, ≥3.8 ng/L). *P*<sub>trend</sub> = P-for-ordinal trend, calculated using the median value in each category.

There were no consistent associations between drinking water PFOA or PFOS with PCa mortality (**Table 4**). However, PFNA ≥3.8 ng/L compared to <2 ng/L was suggestively associated with higher PCa mortality (aHR: 1.22, 95% CI: 0.98, 1.50, *P*_trend_ = 0.15). In analyses using five-year lags, higher concentrations of PFOA (aHR_Q5vsQ1_: 1.10, 95% CI: 0.93, 1.32, *P*_trend_ <.001) and PFOS (aHR_Q5vsQ1_: 1.24, 95% CI: 1.05, 1.48, *P*_trend_ <.001) were associated with higher PCa mortality, though analyses using quintiles revealed significantly lower mortality among those in quintile 3 for PFOA (aHR_Q3vsQ1_: 0.65, 95% CI: 0.54, 0.78) and PFOS (aHR_Q3vsQ1_: 0.70, 95% CI: 0.58, 0.84). Although there was higher PCa mortality with PFNA, estimates were imprecise. Results were similar after applying IPCW (**Supplementary Table 8**).

**Table 4.** Associations of drinking water PFOA, PFOS and PFNA with prostate cancer mortality in New Jersey men with prostate cancer.

|  | **Quintiles of Exposure** |  |  |  |  |  |  |
|---|---|---|---|---|---|---|---|
|  | **Continuous<sup>a</sup>** | **Quintile 1** | **Quintile 2** | **Quintile 3** | **Quintile 4** | **Quintile 5** | ***P*<sub>trend</sub>** |
|  | **aHR (95% CI)** | **aHR (95% CI)** | **aHR (95% CI)** | **aHR (95% CI)** | **aHR (95% CI)** | **aHR (95% CI)** |  |
| **Primary (n=51,289)** |  |  |  |  |  |  |  |
| PFOA |  |  |  |  |  |  |  |
| Cases/Person-years | 3,778/466,843 | 745/93,535 | 741/93,147 | 793/92,964 | 720/93,815 | 779/93,382 |  |
| Minimal<sup>b</sup> | 1.01 (0.98, 1.03) | Ref | 1.01 (0.91, 1.12) | 1.09 (0.98, 1.20) | 0.98 (0.89, 1.09) | 1.02 (0.92, 1.13) | 0.83 |
| Full<sup>c</sup> | 1.01 (0.98, 1.04) | Ref | 0.96 (0.87, 1.07) | 1.07 (0.96, 1.19) | 0.94 (0.84, 1.05) | 0.98 (0.88, 1.10) | 0.58 |
| PFOS |  |  |  |  |  |  |  |
| Cases/Person-years | 3,778/466,843 | 773/93,431 | 766/93,522 | 700/93,494 | 738/93,106 | 801/93,290 |  |
| Minimal<sup>b</sup> | 1.00 (0.99, 1.02) | Ref | 1.01 (0.91, 1.11) | 0.93 (0.83, 1.03) | 1.00 (0.90, 1.11) | 1.05 (0.95, 1.17) | 0.081 |
| Full<sup>c</sup> | 1.00 (0.98, 1.03) | Ref | 1.00 (0.90, 1.10) | 0.95 (0.86, 1.06) | 0.95 (0.85, 1.07) | 1.03 (0.92, 1.15) | 0.29 |
| PFNA<sup>d</sup> |  |  |  |  |  |  |  |
| Cases/Person-years | 3,778/466,843 | 3,726/463,111 | 26/1,933 | 26/1,800 | - | - |  |
| Minimal<sup>b</sup> | 1.00 (1.00, 1.00) | Ref | 1.68 (1.14, 2.47) | 1.65 (1.12, 2.42) | - | - | <.001 |
| Full<sup>c</sup> | 1.00 (1.00, 1.00) | Ref | 1.17 (0.79, 1.73) | 1.42 (0.96, 2.10) | - | - | 0.059 |
| **5-year lag (n=24,840)** |  |  |  |  |  |  |  |
| PFOA |  |  |  |  |  |  |  |
| Cases/Person-years | 1,484/190,028 | 317/38,338 | 296/38,310 | 211/38,029 | 317/37,648 | 343/37,702 |  |
| Minimal<sup>b</sup> | 1.04 (1.00, 1.07) | Ref | 0.94 (0.80, 1.10) | 0.68 (0.57, 0.81) | 0.99 (0.84, 1.15) | 1.07 (0.92, 1.25) | 0.004 |
| Full<sup>c</sup> | 1.04 (1.01, 1.08) | Ref | 0.94 (0.80, 1.11) | 0.65 (0.54, 0.78) | 1.02 (0.86, 1.21) | 1.10 (0.93, 1.32) | 0.001 |
| PFOS |  |  |  |  |  |  |  |
| Cases/Person-years | 1,484/190,028 | 288/38,559 | 322/38,430 | 217/38,041 | 291/37,973 | 366/37,024 |  |
| Minimal<sup>b</sup> | 1.01 (1.00, 1.02) | Ref | 1.13 (0.96, 1.33) | 0.76 (0.64, 0.91) | 1.01 (0.85, 1.19) | 1.27 (1.09, 1.49) | <.001 |
| Full<sup>c</sup> | 1.01 (1.00, 1.02) | Ref | 1.10 (0.94, 1.30) | 0.70 (0.58, 0.84) | 1.02 (0.86, 1.21) | 1.24 (1.05, 1.48) | <.001 |
| PFNA<sup>d</sup> |  |  |  |  |  |  |  |
| Cases/Person-years | 1,484/190,028 | 1,679/188,943 | <11<sup>e</sup>/556 | <11<sup>e</sup>/529 | - | - |  |
| Minimal<sup>b</sup> | 1.00 (1.00, 1.00) | Ref | 0.99 (0.37, 2.65) | 2.17 (1.13, 4.20) | - | - | 0.031 |
| Full<sup>c</sup> | 1.00 (1.00, 1.00) | Ref | 0.78 (0.29, 2.09) | 1.45 (0.74, 2.86) | - | - | 0.39 |

<sup>a</sup>Per interquartile range, <sup>b</sup>Adjusted for age, diagnosis year, <sup>c</sup>Adjusted for <sup>b</sup> and race/ethnicity, marital status, insurance status, stage, treatment (surgery and or radiation), PSA, nSES quintile, NO<sub>2</sub>, PM<sub>2.5</sub>, and NDVI (green space), <sup>d</sup>Due to skewness of PFNA measure, we used categorical cutoffs (<2 ng/L, 2-<3.8 ng/L, ≥3.8 ng/L). <sup>e</sup>Suppressed per registry guidelines. *P*<sub>trend</sub> = P-for-ordinal trend, calculated using the median value in each category.

Higher PFOA exposure was associated with higher CVD mortality (**Table 5**) (aHR_Q2vsQ1_: 1.10, 95% CI: 1.00, 1.20; aHR_Q3vsQ1_: 1.04, 95% CI: 0.94, 1.14; aHR_Q4vsQ1_: 1.05, 95% CI: 0.95, 1.16; aHR_Q5vsQ1_: 1.12, 95% CI: 1.01, 1.24, *P*_trend_ = 0.10). There were no consistent associations between either PFOS or PFNA with CVD mortality. In analyses using five-year lags, higher concentrations of PFOA (aHR_Q5vsQ1_: 1.26, 95% CI: 1.07, 1.48, *P*_trend_ <.001) and PFOS (aHR_Q5vsQ1_: 1.20, 95% CI: 1.02, 1.41, *P*_trend_ = 0.004) were associated with higher PCa mortality, though analyses using quintiles revealed significantly lower mortality among those in quintile 3 for PFOA (aHR_Q3vsQ1_: 0.65, 95% CI: 0.54, 0.78) and PFOS (aHR_Q3vsQ1_: 0.70, 95% CI: 0.58, 0.84). Higher PFNA was associated with lower CVD mortality, but estimates were imprecise. CVD mortality associations with PFAS were similar after applying IPCW to account for competing risks (**Supplementary Table 9**).

**Table 5.** Associations of drinking water PFOA, PFOS and PFNA with cardiovascular mortality in New Jersey men with prostate cancer.

|  |  | Quintiles of Exposure |  |  |  |  |  |
|---|---|---|---|---|---|---|---|
|  | **Continuous<sup>a</sup>** | **Quintile 1** | **Quintile 2** | **Quintile 3** | **Quintile 4** | **Quintile 5** | ***P*<sub>trend</sub>** |
|  | **aHR (95% CI)** | **aHR (95% CI)** | **aHR (95% CI)** | **aHR (95% CI)** | **aHR (95% CI)** | **aHR (95% CI)** |  |
| **Primary (n=51,289)** |  |  |  |  |  |  |  |
| PFOA |  |  |  |  |  |  |  |
| Cases/Person-years | 4,972/466,843 | 879/93,535 | 1,004/93,147 | 1,051/92,964 | 1,011/93,815 | 1,027/93,382 |  |
| Minimal<sup>b</sup> | 0.99 (0.97, 1.01) | Ref | 1.09 (0.99, 1.19) | 0.98 (0.90, 1.08) | 1.01 (0.92, 1.10) | 1.02 (0.93, 1.12) | 0.79 |
| Full<sup>c</sup> | 1.01 (0.99, 1.04) | Ref | 1.10 (1.00, 1.20) | 1.04 (0.94, 1.14) | 1.05 (0.95, 1.16) | 1.12 (1.01, 1.24) | 0.095 |
| PFOS |  |  |  |  |  |  |  |
| Cases/Person-years | 4,972/466,843 | 849/93,431 | 888/93,522 | 984/93,494 | 1,152/93,106 | 1,099/93,290 |  |
| Minimal<sup>b</sup> | 0.99 (0.97, 1.00) | Ref | 0.95 (0.87, 1.05) | 0.94 (0.85, 1.03) | 0.99 (0.90, 1.09) | 0.92 (0.84, 1.02) | 0.24 |
| Full<sup>c</sup> | 1.01 (0.99, 1.02) | Ref | 1.00 (0.90, 1.10) | 0.99 (0.90, 1.09) | 1.04 (0.94, 1.15) | 1.03 (0.93, 1.14) | 0.45 |
| PFNA<sup>d</sup> |  |  |  |  |  |  |  |
| Cases/Person-years | 4,972/466,843 | 4,935/463,111 | 20/1,933 | 17/1,800 | - | - |  |
| Minimal<sup>b</sup> | 1.00 (1.00, 1.00) | Ref | 1.35 (0.87, 2.09) | 1.14 (0.71, 1.84) | - | - | 0.31 |
| Full<sup>c</sup> | 1.00 (1.00, 1.00) | Ref | 1.07 (0.69, 1.67) | 0.93 (0.58, 1.51) | - | - | 0.88 |
| **5-year lag (n=24,840)** |  |  |  |  |  |  |  |
| PFOA |  |  |  |  |  |  |  |
| Cases/Person-years | 1,684/190,028 | 317/38,338 | 369/38,310 | 211/38,029 | 370/37,648 | 417/37,702 |  |
| Minimal<sup>b</sup> | 1.04 (1.01, 1.08) | Ref | 1.16 (1.00, 1.35) | 0.71 (0.60, 0.84) | 1.03 (0.88, 1.19) | 1.14 (0.98, 1.32) | 0.015 |
| Full<sup>c</sup> | 1.06 (1.03, 1.09) | Ref | 1.18 (1.01, 1.37) | 0.71 (0.60, 0.85) | 1.12 (0.95, 1.31) | 1.26 (1.07, 1.48) | <.001 |
| PFOS |  |  |  |  |  |  |  |
| Cases/Person-years | 1,684/190,028 | 301/38,559 | 312/38,430 | 267/38,041 | 368/37,973 | 436/37,024 |  |
| Minimal<sup>b</sup> | 1.01 (1.00, 1.02) | Ref | 0.97 (0.83, 1.14) | 0.89 (0.76, 1.05) | 1.00 (0.86, 1.17) | 1.09 (0.94, 1.27) | 0.039 |
| Full<sup>c</sup> | 1.01 (1.01, 1.02) | Ref | 1.02 (0.87, 1.20) | 0.91 (0.77, 1.08) | 1.09 (0.93, 1.28) | 1.20 (1.02, 1.41) | 0.004 |
| PFNA<sup>d</sup> |  |  |  |  |  |  |  |
| Cases/Person-years | 1,684/190,028 | 1,471/188,943 | <11<sup>e</sup>/556 | <11<sup>e</sup>/529 | - | - |  |
| Minimal<sup>b</sup> | 1.00 (1.00, 1.00) | Ref | 0.62 (0.16, 2.50) | 1.05 (0.34, 3.25) | - | - | 0.98 |
| Full<sup>c</sup> | 1.00 (1.00, 1.00) | Ref | 0.51 (0.13, 2.03) | 0.81 (0.26, 2.52) | - | - | 0.48 |

<sup>a</sup>Per interquartile range, <sup>b</sup>Adjusted for age, diagnosis year, <sup>c</sup>Adjusted for <sup>b</sup> and race/ethnicity, marital status, insurance status, stage, treatment (surgery and or radiation), PSA,  nSES quintile, NO<sub>2</sub>, PM<sub>2.5</sub>, and NDVI (green space), <sup>d</sup>Due to skewness of PFNA measure, we used categorical cutoffs (<2 ng/L, 2-<3.8 ng/L, ≥3.8 ng/L). <sup>e</sup>Suppressed per registry guidelines. *P*<sub>trend</sub> = P-for-ordinal trend, calculated using the median value in each category.

### Mixtures and effect modification

Spearman’s correlations between modeled PFOA and PFOS was very high (r=0.75), with more modest cross-correlations between other PFAS (**Supplementary Figure 2**). Using quantile g-computation, a one-quintile increase across PFOA, PFOS and PFNA was not associated with all-cause mortality (aHR: 1.01, 95% CI: 0.99, 1.03), PCa mortality (aHR: 1.02, 95% CI: 0.97, 1.07) or CVD mortality (aHR: 1.02, 95% CI: 0.98, 1.06) (**Supplementary Table 10**). Although the joint effect across PFOA, PFOS and PFNA were not significantly associated with mortality endpoints, we observed differences in the direction of weights, with PFOS producing strong negative impacts for associations with all-cause and CVD mortality, and PFOA producing negative impacts for associations with PCa mortality (**Supplementary Figure 9**). We observed no evidence of effect modification by age, race, or stage for any PFAS or outcome (**Supplementary Table 11**).

## Discussion

In this population-based cohort study of drinking water PFAS exposure in men with PCa, we found that, for certain analytes, men with PCa residing in areas served by CWS with higher PFAS had higher mortality. The most robust association was between PFOA and CVD mortality, showing a 12% higher CVD mortality rate among those in the highest vs lowest quintile, with suggestive associations between PFOA exposure and all-cause mortality at lower quintiles of exposure. Men exposed to PFNA levels ≥3.8 ng/L had suggestively higher all-cause mortality and PCa mortality compared to those with <2 ng/L. Analyses using five-year lagged PFAS exposure revealed stronger associations between PFOA and PFOS and all-cause and CVD mortality at higher quintiles of exposure.

Earlier epidemiologic studies of PFAS and cancer mortality have yielded inconclusive results. Studies in occupational cohorts that were highly exposed to PFOA from the 1990s found some evidence of higher PCa mortality risk associated with PFOA exposure, but these were based on very small case numbers (<10 events)^74^. As sample sizes increased, with extensive confounding control for occupational, behavioral, and clinical risk factors became more extensive, results have been more equivocal^75^. A 2018 cohort study of 688 PCa survivors conducted within the Danish Diet, Cancer and Health cohort reported no clear associations between serum PFOA or PFOS and PCa survival^76^. Our study adds to this literature with a large sample size of over 50,000 individuals with PCa and assessments of historical drinking water PFOA, PFOS and PFNA.

Our historical reconstruction of PFAS over a 19-year period builds on evidence from prior studies using geospatial exposure assessment to study associations with cancer risk and outcomes^32,77,78^. While spatially-derived exposure measures can introduce measurement error, they are less susceptible to reverse causation bias which can arise using serum internal dose measures when the exposure is affected by underlying disease processes^79^. Earlier studies of geospatial exposure assessment compared populations residing in districts with contamination to those residing in districts without contamination in the same state or province^29,80,81^. More recent studies of cancer incidence and prevalence in the US have relied on federal UCMR3 and UCMR5 data, which cannot easily be compared over time due to differences in the types of water systems and sizes of population served^32,77^. Given that cancer outcomes develop over long periods of time, and the high limits of detection for UCMR3, it has been challenging to integrate data from both databases to study associations of PFAS with cancer outcomes. By leveraging the historic time series of PFAS monitoring results offered by NJDEP monitoring studies and water systems reporting after enactment of safe drinking water regulations, our time-varying exposure model accounted for changes in drinking water PFAS concentrations over time and could be linked to residential histories from NJSCR. Like earlier work leveraging machine learning models to predict drinking water pollutant concentrations, we found excellent discrimination for detect vs non-detect and satisfactory but less accurate predictions of continuous concentrations of PFAS^62^. Model performance suffered when attempting to extrapolate from later to earlier time periods, and from urban to rural settings, suggesting that extrapolation of predicted PFAS to other settings will require retraining and recalculating model parameters. Further modeling refinements could lead to predictions within smaller geographic areas and validation against tap water and well water PFAS results, as well as inclusion of other geospatial predictors that capture variation in PFAS drinking water contamination.

Our study has some limitations. The CWS geographic boundaries cover areas of variable size, which introduces measurement error. The CWS boundary was selected because monitoring results are reported at this level; therefore finer geographic resolution was not available. We expect measurement error to be non-differential with respect to PCa because the PFAS model and input data sources are independent of the NJSCR registry^79^, thereby likely leading to weaker associations than those obtained with more precise exposure assessment. In addition, our models only reflect CWS water systems PFAS exposures and do not capture private well PFAS concentrations, which serve 10% of the NJ population and may have higher PFAS levels due to less frequent testing^82,83^. Our study focuses on legacy contaminants, which may not reflect current exposures to replacement PFAS and related health effects. Lastly, potential residual confounding by individual-level factors, such as healthcare access barriers, water consumption and dietary factors remains a threat to validity. However, our ability to account for time-varying residential neighborhood measures allows us to mitigate major sources of spatial confounding.

In summary, our cohort study of historically constructed drinking water PFAS in men with PCa found that men served by water systems with higher drinking water PFOA levels had higher CVD mortality. There were suggestive associations between drinking water PFOA and all-cause mortality, and between PFNA and PCa-specific mortality, and the magnitude of associations between PFAS and mortality was stronger in analyses using five-year lags. Replication in prospective cohorts with internal dose PFAS measures, assessment of newer replacement short-chain PFAS compounds, and control for additional behavioral risk factors and healthcare access barriers are needed to validate findings.

## Data Availability

Human epidemiologic data for this study were requested from the New Jersey State Cancer Registry for the purposes of this research, and Institutional Review Board and Data Use Agreements between Rutgers, Dana-Farber, and the New Jersey State Cancer Registry prohibit sharing of these data outside of the research team. However, the analytic code and drinking water per- and polyfluoroalkyl substances exposure models used for this study are available on GitHub (PROVIDE LINK). Inquiries can be directed to

## Notes

**Funding:** HSI was supported by Prostate Cancer Research, UK, NIEHS K01ES035734 and pilot grants from NIEHS P30ES005022 and NCI P30CA072720. JEH was supported by NIEHS P30ES000002. This work was supported by the National Cancer Institute’s Surveillance, Epidemiology, and End Results (SEER) Program (75N91021D00009) and the Centers for Disease Control and Prevention (CDC) National Program of Cancer Registries (5NU58DP007117) with additional support from the State of New Jersey and the Rutgers Cancer Institute of New Jersey. The content is solely the responsibility of the authors and does not necessarily represent the official views of the National Institutes of Health.

### Competing Interest Statement

The authors have declared no competing interest.

### Author Declarations

The institutional review boards of the Dana-Farber Cancer Institute and Rutgers University, The State University, gave ethical approval for this work and determined that, because existing data sources were used, no written consent was required for participation in the study.

