## Supplementary Material for "Associations of drinking water per- and polyfluoroalkyl substances with cause-specific mortality in men with prostate cancer"

**Associations of historically reconstructed drinking water per- and polyfluoroalkyl substances with cause-specific mortality in New Jersey men with prostate cancer**

Hari S. Iyer, Megan R. Shanahan, Stefanie Joseph, Julianne Varga, Mike Z. He, Saum Ghodoussipour, Antoinette M. Stroup, Gerald Harris, Panos Georgopoulos, Robert Laumbach, Veronica M. Vieira, Jaime E. Hart, Emily S. Barrett, Elisa V. Bandera

**Supplementary methods.**

**Supplementary Table 1. Results, limits of detection, and summary statistics for number of results per Community Water System for PFOA, PFOS and PFNA, New Jersey, 2006-2025**

**Supplementary Table 2. Predictors for historical reconstruction drinking water PFAS model, New Jersey, 2006-2025**

**Supplementary Table 3. Data Axle Business Locations, 2006-2024**

**Supplementary Table 4: Hyperparameters selected for final XGBoost Community Water Systems PFAS model, New Jersey, 2006-2025**

**Supplementary Table 5. Performance statistics for XGBoost Community Water Systems PFAS model, New Jersey, 2006-2025**

**Supplementary Table 6. Characteristics of New Jersey Men with Prostate Cancer Stratified by Quintile of PFOS**

**Supplementary Table 7. Characteristics of New Jersey Men with Prostate Cancer Stratified by Quintile of PFNA**

**Supplementary Table 8. Associations of drinking water PFOA, PFOS and PFNA with prostate cancer mortality in New Jersey men with prostate cancer using inverse probability of censoring weights for competing risks**

**Supplementary Table 9. Associations of drinking water PFOA, PFOS and PFNA with cardiovascular disease mortality in New Jersey men with prostate cancer using inverse probability of censoring weights for competing risks**

**Supplementary Table 10. Quantile g-computation results for mixtures of PFOA, PFOS and PFNA quintiles and all-cause, prostate cancer, and cardiovascular disease mortality in men with prostate cancer, New Jersey, 2006-2024.**

**Supplementary Table 11. Associations of PFOA with CVD mortality in men with prostate cancer, stratified by race and ethnicity, age and state at diagnosis, New Jersey, 2006-2024.**

**Supplementary Figure 1. Participant CONSORT Diagram**

**Supplementary Figure 2. Correlation matrix for PFAS and neighborhood contextual factors and pollutants**

**Supplementary Figure 3. XGBoost Model for PFOA, Feature Importance**

**Supplementary Figure 4. XGBoost Model for PFOS, Feature Importance**

**Supplementary Figure 5. XGBoost Model for PFNA, Feature Importance**

**Supplementary Figure 6. Maps of predicted PFOA concentrations across 566 Community Water Systems, New Jersey, 2006-2025**

**Supplementary Figure 7. Maps of predicted PFOS concentrations across 566 Community Water Systems, New Jersey, 2006-2025**

**Supplementary Figure 8. Maps of predicted PFNA concentrations across 566 Community Water Systems, New Jersey, 2006-2025**

**Supplementary Methods**

*1. Residential history data linkage*

Residential histories were constructed in two major steps. In the first step, a list of addresses were compiled through a linkage with LexisNexis, a commercial credit reporting company and data provider that uses real estate/tax assessor records, deed transfers and mortgage records, vehicle registrations, driver’s license records, court filings, professional licenses, voter registrations, death records, marriage and divorce record from certain states, criminal history records and business records^1^. Lexis Nexis applied a probabilistic matching algorithm to the New Jersey State Cancer Registry PCa cases based on name, date of birth, Social Security Number, and geographic coordinates for date of diagnosis to generate a unique personal identifier that can be used to merge back to the New Jersey State Cancer Registry data^2^.

Upon receiving the Lexis Nexis file, which contains a unique personal identifier, the address street, city, state, ZIP code, latitude, longitude, geographic accuracy, and date of start and date of end at the address, individuals from the New Jersey State Cancer Registry applied the Match*Pro probabilistic algorithm to prepare a list of addresses for each individual patient with PCa for this study. Prior to running the Match * Pro algorithm, the registry officials cleaned dates by scaling them to months before or after diagnosis, checking for inconsistent city, state and ZIP codes, as well as unique identifiers, excluding participants who are missing date of diagnosis. Duplicate addresses are removed using the SAS SORT function. The Match*Pro algorithm then applies a probabilistic match score using the SAS SPEDIS, SOUNDEX and COMPARE functions using the unique identifiers and address information with information about priority (proximity to date of diagnosis) and based on the confidence in the matches. This confidence is weighted by the street name threshold parameters and distance between addresses from the cancer registry file and LexisNexis file. A step of manual review was then performed to review borderline matches, name discrepancies, and distance discrepancies. Finally, gaps in address histories were resolved by extending the stop date of an old address to match the start date of a new address in the history. Further details are available elsewhere^3^.

A total of 427,201 addresses resulted from the Lexis Nexis probabilistic matching and further cleaning by New Jersey State Cancer Registry data analysts. Of these, 82% were geocoded to the address-level, 10% were geocoded to the street address level, and 2% were joined to the nearest intersection, 9 digit ZIP code or street name. Participants stayed a mean of 9.8 (SD=11.4) years at a given address, with a mean of 1.7 (SD=1.1) addresses per person. The number of addresses per individual ranged from 1-22. Of all of the addresses identified by LexisNexis, 82% were in New Jersey, with the remaining 18% capturing addresses from remaining states and US territories. Because our PFAS model was only available in New Jersey, we restricted the study population to those with complete address histories in the state of New Jersey.

*2. PFAS drinking water exposure model: Overview*

NJ is one of the first states to conduct statewide monitoring and surveillance of PFAS in the drinking water, with the earliest results obtained in 2006. We sought to leverage this historical database to generate PFOA, PFOS and PFNA predictions at Community Water Systems (CWS) level for the state of NJ over 3-month intervals from January 2006 to December 2025. These three analytes were selected because they have been monitored for the longest period of time in NJ^4^, and were the subject of regulations between 2018 to 2020^5^. CWS are public water systems that supply piped drinking water to the same population of year-round residents, serving at least 15 service connections used by year-round residents or regularly serving at least 25 year-round residents. In NJ, there are 566 CWS, serving an estimated 87% of the total population^6^.

The CWS geographic unit was selected because PFAS results are reported by CWS and finer scale geographic assessment was not possible, and because the US Environmental Protection Agency (EPA) has created polygon shapefiles to capture CWS boundaries in NJ and other states. The EPA boundaries were developed using random forest models based on key variables (imperviousness, 2020 housing unit density area, % public use in 1990, % sewer use in 1990, 1990 housing units, % housing unit change 1990-2020, distance to public water intake) and aligned with census block group boundaries to allow them to be superimposed on top of existing census administrative units. More details regarding the boundary creation are available elsewhere^7^.

In review of the boundaries and PFAS monitoring results, we identified one water system boundary that had changed IDs over time. We therefore manually created a polygon to capture census tracts served by that boundary and added it to the EPA CWS database to obtain our final number of 566 CWS boundaries.

*2.1. PFAS data sources*

Data for PFOA, PFOS and PFNA results in drinking water were obtained from two data sources. The first are the reporting results from surveillance and monitoring studies conducted as part of NJ’s early monitoring studies from 2006-2013^8,9^. These data were accessed through contacts at the New Jersey Department of Environmental Protection (NJDEP), who released deidentified results from monitoring studies between 2006 and 2018. In brief, sample locations for the monitoring studies were first selected based on probable contamination, and then selected to provide sufficient characterization of state-wide patterns. The second set of data were accessed as part of regulatory reporting by water systems from 2018-2025 accessed from the New Jersey Drinking Water Watch database^10^. We excluded unfinished results and those with documented quality control issues (n=1640). All results were converted to ng/L units. There were 1,456 results retained from the NJDEP monitoring studies, and 14,935 results retained from the routine monitoring data after MCLs were implemented resulting in 16,391 results covering 80 CWS. Further details about these sources, including the assay methods and limits of detection, were described previously^5^.

In addition to NJ state drinking water reporting results, we obtained data from the Unregulated Contaminant Monitoring Rule (UCMR), conducted between 2013-2015 (UCMR3) and 2023-2026 (UCMR5). The UCMR3 collected results from Public Water Systems (PWS), a broader category that encompasses CWS and smaller water systems with piped water to at least 15 service connections or ≥25 people for at last 60 days each year, serving >10,000 individuals and required reporting of PFOA, PFOS, and PFNA. However, the Minimum Reporting Levels for PFOA (20 ng/L), PFOS (40 ng/L) and PFNA (20 ng/L) were substantially higher than the limits of detection for the NJDEP studies and routine reporting data. There were 4,383 results covering 175 PWS available in the UCMR3. For UCMR5, there were lower Minimum Reporting Levels for PFOA (4 ng/L), PFOS (4 ng/L) and PFNA (4 ng/L) (**Supplementary Table 1**). There were 5,387 observations covering 263 PWS. The UCMR3 used EPA Method 537 Rev 1.1, while UCMR5 used EPA Method 533 for the three key PFAS assessments. More details are available elsewhere^11–14^.

*2.3. Predictors*

We chose a total of 67 predictors based on prior evidence of associations between spatial predictors and PFAS contamination^13,15–18^ (**Supplementary Table 2**). Key sources that have previously been associated with presence of drinking water PFAS contamination included airports, military bases and training facilities, businesses with evidence of PFAS manufacturing, and firefighting stations. For water and hydrological parameters, we reviewed earlier studies that have generated groundwater PFAS predictions based on data from the United States Geological Survey^18^. In addition, we used sociodemographic characteristics of the population and outdoor air pollution as potential predictors of industrial activity or occupational activities that may be linked to greater PFAS exposure^13^.

Data on military bases and training facilities were obtained from the Military Installations, Ranges and Training Areas (MIRTA)^19^ and the Formerly Used Defense Sites (FUDS)^20^ databases. The FUDS database is used to guide environmental cleanup following military use of land. We identified 50 individual locations within NJ across FUDS (n=42) and MIRTA (n=8). Data on airports were obtained from the New Jersey Department of Transportation (NJDOT) (n=41) database, which reflected active airports in NJ in 2021.

Data on firefighting stations were obtained from public databases of address locations^21,22^. These websites were identified through searches of stations augmented by ChatGPT, and data were downloaded in February 2026. We used ChatGPT to combine both sources and to standardize the format of the addresses for geoprocessing, and to identify initial duplicates between sources. A subset of locations were verified manually using Google Maps. Once sources were cleaned, we geocoded the addresses in ArcGIS Pro, sorted by latitude and longitude, and dropped observations within 500m of each other as duplicates. We identified 897 fire stations across NJ.

Data on business locations were obtained from the Data Axle business history database, which captures business locations annually from 1999 to 2024^23^. Although this database is primarily used for marketing, the data curators employ data quality and consistency checks by confirming businesses are operating during that year and confirming accuracy of geocodes^24^. We used North American Industry Classification Codes (NAICS) to filter businesses that have been previously associated with PFAS contamination^15^. In order to capture changes in business type and geographic spread over time, we filtered the business histories to preserve cross-sectional location data in 2000, 2005, 2010, 2015, and 2020. Numbers of locations by business type are provided in **Supplementary Table 3**.

In order to incorporate hydrological features as predictors, we obtained processed hydrological geospatial data on average percentage of clay, well depth, depth to water, estimated nitrogen loading from septic systems, and groundwater recharge, as well as distance to other major PFAS sources previously processed and used by Tokranov and colleagues in a study to predict nation-wide groundwater concentrations of 24 PFAS^18^. These geospatial features were processed as rasters to generate point locations when input data were originally 800m resolution, or by applying focal statistics within 500m radial buffers, except for well depth which was processed at 250m resolution and extracted to point locations. Output rasters were provided at 1km resolution, the scale used in this study.

Sociodemographic context was estimated from US Census data in 2000 and 2010 at the Census Tract level. We obtained measures of population density, and educational attainment, poverty, racial composition, household income, and household wealth based on prior work^25,26^. Measures were obtained for 2000 and 2010 to enable time-varying predictions.

Outdoor air pollution may indirectly reflect industrial sources of pollution and therefore may predict higher propensity for drinking water contamination. We therefore obtained publicly available fine particulate matter (PM_2.5_) and nitrogen dioxide (NO_2_) predictions from the Schwartz laboratory at the Harvard T. H. Chan School of Public Health, which were produced using ensemble models from government monitoring databases and academic research studies from 2156 locations and 912 locations for PM2.5 and NO2 respectively across the US^27,28^. Predictors included geocodes, year, elevation, meteorologic parameters, and satellite data on vegetation, outdoor light at night, and carbon emissions. The models displayed 10-fold cross-validated R^2^ values of 0.89 for annual predictions of PM_2.5_ and values of 0.79 for annual predictions of NO_2_. We obtained annual estimates from 2000 to 2016 at 1km resolution, to enable time-varying predictions.

In addition to geospatial sources of PFAS, we added a variable to indicate the season as a predictor in our PFAS models (January-March, April-June, July-September, October-December) given evidence of seasonal variation in PFAS levels from earlier work^5^. We chose not to include year given the strong time trends that have been previously reported and to allow potential for extrapolating predictions to other time periods and geographic areas.

*2.4 Predictor data processing*

After collecting all databases, we first restricted the nationwide datasets to the state boundaries of New Jersey using clip tools in ArcGIS Pro. For point locations, we calculated inverse distance metrics from the centroid of census block groups in New Jersey to each type of PFAS contamination source location (airports, military sites, firefighting stations, businesses). We varied the distance radius (1km, 3km and 5km) and the number of locations (5, 10, and 15) using the “generate near table” tool in ArcGIS Pro, setting “input” features as the census block group centroids and the “near” features as the PFAS contamination sources. We then used zonal statistics to average these inverse distance measures within the New Jersey CWS polygon boundaries to obtain a CWS-level estimate of the inverse distance measure for each contamination source type.

For socioeconomic data from the census at census tract level, we calculated zonal statistics for each CWS, averaging each SES measure within that CWS boundary. For raster data, we calculated zonal statistics for average PM2.5, NO2 and each of the hydrological parameters within each CWS boundary.

For certain data, we had time-varying data (business locations: 2000, 2005, 2010, 2015, 2020; census data: 2000, 2010; air pollution: 2000-2016 per year). In these cases, we preserved the temporal resolution of these databases when joining data.

Once all PFAS source measures and predictors were linked to CWS boundaries, we prepared an input database organized by year (2006-2025) and by quarter (January-March, April-June, July-September, October-December) to allow for quarterly estimates to be modeled. In earlier work, we found evidence of season patterns in certain PFAS analytes in water monitoring data^5^.

*2.5. Model validation*

We then fit two-phase XGBoost models using 10-fold cross-validation using a 70%/30% training/test split in our primary analysis following guidance for machine learning-based drinking water contaminant prediction^29^. XGBoost models were selected because these have been used in previous efforts to model groundwater concentrations of PFAS across the contiguous US^18^, and have been shown to produce good performance in reviews of drinking water exposure modeling^29^. In the first phase, we predicted detectable vs non-detectable concentrations using a logistic regression classifier. In the second phase, we predicted average concentrations using a linear regression classifier. Hyperparameters were tuned using grid searches within the xgboost package in R, first to optimize nrounds, eta and max_depth, next to optimize colsample_bytree and subsample, and lastly to optimize gamma, lambda, and alpha hyperparameters. The final set of hyperparameters used for each model is provided in **Supplementary Table 4**.

In order to evaluate performance of the XGBoost models, we presented Area Under the Curve (AUC) for the detect vs non-detect results, and the Root Mean Squared Error (RMSE), and R^2^ (the variation in PFAS explained by predictors) for concentrations. These metrics were chosen to assess discrimination of the model for identifying detectable PFAS, and to evaluate the performance of models to estimate concentrations across water systems. Results for test vs training data are presented in **Supplementary Table 5**. Training AUC for detect vs non-detect PFOA (0.94), PFOS (0.94), and PFNA (0.95) were excellent, and declined slightly in test samples (PFOA: 0.91, PFOS: 0.90, PFNA: 0.92). For PFAS concentrations, R^2^ were generally acceptable (PFOA: 0.75, PFOS: 0.64, PFNA: 0.88). These R^2^ declined in test samples (PFOA: 0.68, PFOS: 0.53, PFNA: 0.75) but remained acceptable.

We compared modeled drinking water PFAS predictions and tap water collection from a pilot study of 71 NJ men with PCa^30^ within the same 3-month period showed moderate correlations for PFOA (r=0.44, p<.001) and PFOS (r=0.33, p <.01) but weaker correlations for PFNA (r=0.03). The weak correlation between tap water and modeled PFNA may have been due to the strong geographic clustering of high PFNA values and strong skew in PFNA drinking water predicted values, leading to few participants in our pilot study residing in the area with high PFNA concentrations.

We checked correlations between our key neighborhood factors and modeled PFAS values (**Supplementary Figure 2**), and found no evidence of potential collinearity. Feature importance for models revealed that PM_2.5_ and NO_2_ were strong predictors for all models (**Supplementary Figure 3, Supplementary Figure 4, Supplementary Figure 5**). Additional predictors that remained consistently high in feature importance included proximity to business locations, season, and proximity to military sites for PFOS and PFNA. For PFOA, hydrological measures had higher feature importance.

We conducted sensitivity analyses using different training test splits based on time period (train: 2018-2025/ test: 2006-2017) and CWS population density (train: ≥1000 people/mi^2^, <1000 people/mi^2^). Performance was poor across validation metrics for detect vs non-detect and for predicted vs observed concentrations when training and test samples were taken from different time periods and different geographic areas (**Supplementary Table 4, Supplementary Table 5**). This suggests that out-of-sample predictions using the weights obtained from our random samples in NJ would not perform well, and so rather than extrapolating from these models, it would be best to repeat this approach in other states or time periods if data are available.

16.	Tokranov AK, LeBlanc DR, Pickard HM, Ruyle BJ, Barber LB, Hull RB, et al. Surface-water/groundwater boundaries affect seasonal PFAS concentrations and PFAA precursor transformations. Environ Sci Process Impacts. 23(12):1893–905. doi:10.1039/d1em00329a PubMed PMID: 34779453; PubMed Central PMCID: PMC8673475.

23.	Data Axle. Historical business files /Data Axle. Rutgers Center for Climate, Health and Healthcare; 2026.

**Supplementary Table 1. Results, limits of detection, and summary statistics for number of results per Community Water System for PFOA, PFOS and PFNA, New Jersey, 2006-2025**

| **PFAS** | **Results (n)** | **Limit of Detection/Minimum Reporting Level** | **Median [IQR] number of results per CWS** | **Range (min, max) of results per CWS** |
| --- | --- | --- | --- | --- |
| PFOA | 8,687 | <0.27 to <10 ng/L  20 ng/L (UCMR3, MRL)  4 ng/L (UCMR5, MRL) | 10 [4, 24] | 1, 621 |
| PFOS | 8,800 | <0.21 to <4 ng/L  40 ng/L (UCMR3, MRL)  4 ng/L (UCMR5, MRL) | 10 [4, 24] | 1, 541 |
| PFNA | 8,674 | <0.20 to <5 ng/L  20 ng/L (UCMR3, MRL)  4 ng/L (UCMR5, MRL) | 10 [4, 24] | 1, 520 |

Abbreviation: LOD=Limit of Detection (NJDEP monitoring studies and voluntary reporting);

MRL=Minimum Reporting Level (UCMR only)

**Supplementary Table 2. Predictors for historical reconstruction drinking water PFAS model, New Jersey, 2006-2025**

| **Predictor** | **Data source** | **Time period** | **Spatial resolution** | **Number of locations (points only)** | **Number of predictors** | **Description** |
| --- | --- | --- | --- | --- | --- | --- |
| Former military bases and training areas | FUDS | 2021 | Point | 42 | 5 | Captures facilities owned by US government under jurisdiction of the Secretary of Defense. Properties were transferred from the Department of Defense prior to October 17, 1986. |
| Military installations, ranges and training areas | MIRTA | 2009 | Points | 8 | 4 | Gives locations of most commonly known US Department of Defense sites from the 2009 Base Structure Report inventory. Installations are commonly referred to as a base, camp, post, station, yard, center, or homeport facility for any ship. |
| Firefighting stations | Firefighting county offices | 2026 | Points | 897 | 6 | Searched for firefighting stations through ChatGPT, which identified ExpertGPS.com and CountyOffice.org as sources. Locations were downloaded in February 2026. A subset of locations were verified through Google Maps searches. |
| Airports | NJDOT | 2021 | Points | 41 | 5 | Provides locations of 41 pubic use airports. |
| Business locations | Data Axle | 2000-2023 | Points | 25,448 | 6 | Provides locations of businesses classified using NAICS codes for industrial categories defined in previous studies of PFAS water contamination^15^ |
| Sociodemographic measures | US Census, American Community Survey | 2000, 2010 | Census tract | Not applicable | 28 | Pulled income, wealth, educational attainment, racial composition, segregation indices at tract-level based on prior evidence for associations between socioeconomic status, racial composition and environmental exposures^25^ |
| Hydrological measures | US Geological Survey, |  | Raster (1km x 1km) |  | 10 | Averaged raster values within the CWS boundary using zonal statistics |
| Fine particulate matter (PM2.5) | Modeled (Schwartz laboratory) | 2000-2016 | Raster (1km x 1km) | Not applicable | 1 | Averaged pixel values within the CWS boundary using zonal statistics |
| Nitrogen dioxide (NO2) | Modeled (Schwartz laboratory) | 2000-2016 | Raster (1km x 1km) | Not applicable | 1 | Averaged pixel values within the CWS boundary using zonal statistics |

Abbreviations: FUDS=Formerly Used Defense Sites, MIRTA=Military Installations, Ranges and Training Areas, NJDOT=New Jersey Department of Transportation

**Supplementary Table 3. Data Axle Business Locations, 2006-2024**

| **NAICS code** | **Business type** | **No. NJ facilities** |
| --- | --- | --- |
| 562213 | Solid Waste Combustors and Incinerators | - |
| 326112 | Plastics Packaging Film and Sheet (including Laminated) Manufacturing | - |
| 323111 | Commercial Printing (except Screen and Books) | 9043 |
| 424690 | Other Chemical and Allied Products Merchant Wholesalers | 2698 |
| 333318 | Other Commercial and Service Industry Machinery Manufacturing | 1331 |
| 323120 | Support Activities for Printing | 1238 |
| 313310 | Textile and Fabric Finishing Mills | 1194 |
| 322220 | Paper Bag and Coated and Treated Paper Manufacturing | 990 |
| 334419 | Other Electronic Component Manufacturing | 912 |
| 332999 | All Other Miscellaneous Fabricated Metal Product Manufacturing | 862 |
| 332812 | Metal Coating, Engraving (except Jewelry and Silverware), and Allied Services to Manufacturing | 792 |
| 325998 | All Other Miscellaneous Chemical Product and Preparation Manufacturing | 780 |
| 562219 | Other Nonhazardous Waste Treatment and Disposal | 620 |
| 324110 | Petroleum Refineries | 543 |
| 335999 | All Other Miscellaneous Electrical Equipment and Component Manufacturing | 466 |
| 325211 | Plastics Material and Resin Manufacturing | 458 |
| 332813 | Electroplating, Plating, Polishing, Anodizing, and Coloring | 422 |
| 334413 | Semiconductor and Related Device Manufacturing | 412 |
| 314910 | Textile Bag and Canvas Mills | 274 |
| 322219 | Other Paperboard Container Manufacturing | 262 |
| 325510 | Paint and Coating Manufacturing | 260 |
| 326113 | Unlaminated Plastics Film and Sheet (except Packaging) Manufacturing | 241 |
| 322121 | Paper (except Newsprint) Mills | 213 |
| 325611 | Soap and Other Detergent Manufacturing | 179 |
| 562211 | Hazardous Waste Treatment and Disposal | 165 |
| 313220 | Narrow Fabric Mills and Schiffli Machine Embroidery | 153 |
| 325612 | Polish and Other Sanitation Good Manufacturing | 144 |
| 325199 | All Other Basic Organic Chemical Manufacturing | 120 |
| 313210 | Broadwoven Fabric Mills | 117 |
| 322130 | Paperboard Mills | 96 |
| 324191 | Petroleum Lubricating Oil and Grease Manufacturing | 85 |
| 314110 | Carpet and Rug Mills | 79 |
| 313320 | Fabric Coating Mills | 70 |
| 562212 | Solid Waste Landfill | 69 |
| 316110 | Leather and Hilde Tanning and Finishing | 69 |
| 313110 | Fiber, Yarn, and Thread Mills | 57 |
| 313230 | Nonwoven Fabric Mills | 29 |
| 562112 | Hazardous Waste Collection | 5 |

**Supplementary Table 4: Hyperparameters selected for final XGBoost Community Water Systems PFAS model, New Jersey, 2006-2025**

| **Train/test split** | **Phase 1 (detect)** | | | **Phase 2 (concentration)** | | |
| --- | --- | --- | --- | --- | --- | --- |
| **Random 70%/30%** |  | | |  | | |
| **Hyperparameters** | **PFOA (bin)** | **PFOS (bin)** | **PFNA (bin)** | **PFOA (reg)** | **PFOS (reg)** | **PFNA (reg)** |
| nounds | 400 | 1000 | 1000 | 300 | 1000 | 1000 |
| eta | 0.08 | 0.05 | 0.04 | 0.03 | 0.04 | 0.03 |
| Gamma | 0.05 | 0 | 0.1 | 0.05 | 0.1 | 0.05 |
| Colsample_bytree | 0.7 | 0.06 | 0.8 | 0.8 | 1 | 1 |
| Min_child_weight | 2 | 2 | 3 | 1 | 1 | 4 |
| Subsample | 1 | 1 | 0.9 | 0.9 | 1 | 0.8 |
| Lambda | 1 | 1 | 1 | 0.5 | 0 | 2 |
| Alpha | 1 | 0 | 0 | 0.5 | 0 | 0.5 |
| Max_depth | 4 | 7 | 4 | 7 | 6 | 6 |
| **Time period (2018-2025, 2006-2017)** |  |  |  |  |  |  |
| Nounds | 400 | 600 | 1000 | 500 | 600 | 1000 |
| Eta | 0.08 | 0.04 | 0.03 | 0.025 | 0.04 | 0.02 |
| Gamma | 0 | 0.1 | 0.1 | 0 | 0.2 | 0.1 |
| Colsample_bytree | 0.7 | 0.8 | 1 | 0.7 | 1 | 1 |
| Min_child_weight | 2 | 3 | 1 | 3 | 5 | 1 |
| Subsample | 0.9 | 0.9 | 0.8 | 0.8 | 0.9 | 0.7 |
| Lambda | 1 | 1 | 0 | 0.5 | 1 | 0 |
| Alpha | 0 | 1 | 0 | 0 | 0 | 0 |
| Max_depth | 5 | 7 | 3 | 6 | 4 | 4 |
| **Water System Population Density/Geography (1000+ people/mi^2^, <1000 people/mi^2^)** |  |  |  |  |  |  |
| nounds | 400 | 800 | 1000 | 250 | 800 | 1000 |
| eta | 0.08 | 0.05 | 0.03 | 0.025 | 0.025 | 0.02 |
| Gamma | 0.01 | 0 | 0.1 | 0 | 0.1 | 0.1 |
| Colsample_bytree | 0.9 | 0.8 | 1 | 0.7 | 0.85 | 1 |
| Min_child_weight | 4 | 3 | 1 | 3 | 1 | 1 |
| Subsample | 1 | 1 | 0.8 | 0.8 | 0.8 | 0.7 |
| Lambda | 1 | 1 | 0 | 0.5 | 0 | 0 |
| Alpha | 1 | 1 | 0 | 0 | 0 | 0 |
| Max_depth | 4 | 5 | 3 | 5 | 6 | 4 |

**Supplementary Table 5. Performance statistics for XGBoost Community Water Systems PFAS model, New Jersey, 2006-2025**

|  | **C-Statistic** | | **R^2^** | | **RMSE** | |
| --- | --- | --- | --- | --- | --- | --- |
| **Scenario (N_train_, N_test_)** | **Train** | **Test** | **Train** | **Test** | **Train** | **Test** |
| **Random 70%/30%** |  |  |  |  |  |  |
| PFOA (6160/2597) | 0.94 | 0.91 | 0.75 | 0.68 | 0.38 | 0.43 |
| PFOS (6072/2559) | 0.94 | 0.90 | 0.63 | 0.53 | 0.40 | 0.47 |
| PFNA (6081/2563) | 0.95 | 0.92 | 0.86 | 0.75 | 0.44 | 0.56 |
| **Time period (2018-2025, 2006-2017)** | | | | | | |
| PFOA (6686/2071) | 0.91 | 0.56 | 0.66 | -1.32 | 0.38 | 1.18 |
| PFOS (6677/1954) | 0.90 | 0.63 | 0.55 | -0.22 | 0.42 | 1.12 |
| PFNA (6875/1769) | 0.93 | 0.82 | 0.79 | -0.13 | 0.45 | 0.87 |
| **Water System Population Density/Geography (≥1000 people/mi^2^, <1000 people/mi^2^)** | | | | | | |
| PFOA (5252/3505) | 0.93 | 0.74 | 0.63 | -0.43 | 0.40 | 0.91 |
| PFOS (5204/3427) | 0.90 | 0.76 | 0.55 | 0.005 | 0.40 | 0.67 |
| PFNA (5200/3444) | 0.95 | 0.60 | 0.79 | -0.25 | 0.55 | 1.15 |

Legend: N_train_ = number of training results, N_test_ = number of test results, PFOA=perfluorooctanoic acid, PFOS=perfluorooctane sulfonic acid, PFNA=perfluorononanoic acid

**Supplementary Table 6. Characteristics of New Jersey Men with Prostate Cancer Stratified by Quintile of PFOS**

|  | **Quintiles of PFOS** | | | | |
| --- | --- | --- | --- | --- | --- |
| **Characteristic (n, %)** | **Quintile 1** | **Quintile 2** | **Quintile 3** | **Quintile 4** | **Quintile 5** |
| **N** | 14662 | 11561 | 9654 | 7373 | 8039 |
| **PFOS, ng/L (median, [IQR])** | 1.01 [1.01, 1.02] | 1.07 [1.04, 1.09] | 1.26 [1.20, 1.34] | 2.10 [1.70, 2.54] | 5.37 [4.34, 8.92] |
| **Age at diagnosis, years (mean, SD)** | 67.08 (9.33) | 66.93 (9.41) | 67.12 (9.42) | 67.23 (9.44) | 67.50 (9.38) |
| **Age at diagnosis, years** |  |  |  |  |  |
| <50 | 510 (3%) | 412 (4%) | 302 (3%) | 277 (4%) | 242 (3%) |
| 50-60 | 3095 (21%) | 2522 (22%) | 2136 (22%) | 1562 (21%) | 1688 (21%) |
| 60-70 | 5816 (40%) | 4628 (40%) | 3738 (39%) | 2839 (39%) | 3108 (39%) |
| 70-80 | 4078 (28%) | 3066 (27%) | 2653 (27%) | 2062 (28%) | 2291 (28%) |
| ≥80 | 1163 (8%) | 933 (8%) | 825 (9%) | 633 (9%) | 710 (9%) |
| **Year of diagnosis** |  |  |  |  |  |
| 2006-2010 | 4469 (30%) | 3178 (27%) | 3017 (31%) | 3538 (48%) | 4500 (56%) |
| 2010-2014 | 6885 (47%) | 4456 (39%) | 2853 (30%) | 1807 (25%) | 567 (7%) |
| 2014 -2017 | 3308 (23%) | 3927 (34%) | 3784 (39%) | 2028 (28%) | 2972 (37%) |
| **Race/Ethnicity** |  |  |  |  |  |
| NHW | 11339 (77%) | 8120 (70%) | 6722 (70%) | 4849 (66%) | 4873 (61%) |
| NHB | 1984 (14%) | 1844 (16%) | 1660 (17%) | 1371 (19%) | 1539 (19%) |
| NHAAPI | 458 (3%) | 456 (4%) | 382 (4%) | 316 (4%) | 365 (5%) |
| Hispanic | 881 (6%) | 1141 (10%) | 890 (9%) | 837 (11%) | 1262 (16%) |
| **Marital Status** |  |  |  |  |  |
| No | 2896 (20%) | 2351 (20%) | 1915 (20%) | 1552 (21%) | 1755 (22%) |
| Yes | 9353 (64%) | 7141 (62%) | 6102 (63%) | 4531 (61%) | 4985 (62%) |
| Missing | 2413 (16%) | 2069 (18%) | 1637 (17%) | 1290 (17%) | 1299 (16%) |
| **Insurance** |  |  |  |  |  |
| Uninsured | 951 (6%) | 791 (7%) | 609 (6%) | 492 (7%) | 614 (8%) |
| Private | 4305 (29%) | 3348 (29%) | 2761 (29%) | 2136 (29%) | 2047 (25%) |
| Medicaid | 185 (1%) | 194 (2%) | 134 (1%) | 106 (1%) | 164 (2%) |
| Medicare | 5915 (40%) | 4160 (36%) | 3589 (37%) | 2794 (38%) | 2856 (36%) |
| Other government | 41 (0%) | 28 (0%) | 28 (0%) | 23 (0%) | 16 (0%) |
| Unknown | 3265 (22%) | 3040 (26%) | 2533 (26%) | 1822 (25%) | 2342 (29%) |
| **Gleason (combined)** |  |  |  |  |  |
| <7 | 7667 (52%) | 6455 (56%) | 5444 (56%) | 4018 (54%) | 4567 (57%) |
| ≥7 | 5735 (39%) | 4083 (35%) | 3373 (35%) | 2736 (37%) | 2956 (37%) |
| Unknown | 1260 (9%) | 1023 (9%) | 837 (9%) | 619 (8%) | 516 (6%) |
| **Stage** |  |  |  |  |  |
| Localized | 11160 (76%) | 8251 (71%) | 6460 (67%) | 4631 (63%) | 4421 (55%) |
| Regional | 1100 (8%) | 811 (7%) | 634 (7%) | 443 (6%) | 354 (4%) |
| Distant | 590 (4%) | 448 (4%) | 344 (4%) | 222 (3%) | 179 (2%) |
| Unknown | 1812 (12%) | 2051 (18%) | 2216 (23%) | 2077 (28%) | 3085 (38%) |
| **Surgery** |  |  |  |  |  |
| No | 9613 (66%) | 7614 (66%) | 6389 (66%) | 4760 (65%) | 5339 (66%) |
| Yes | 4849 (33%) | 3758 (33%) | 3103 (32%) | 2393 (32%) | 2510 (31%) |
| Unknown | 200 (1%) | 189 (2%) | 162 (2%) | 220 (3%) | 190 (2%) |
| **Hormone treatment** |  |  |  |  |  |
| No | 10586 (72%) | 8406 (73%) | 6852 (71%) | 5049 (68%) | 5335 (66%) |
| Yes | 3799 (26%) | 2905 (25%) | 2569 (27%) | 2034 (28%) | 2434 (30%) |
| Unknown | 277 (2%) | 250 (2%) | 233 (2%) | 290 (4%) | 270 (3%) |
| **Radiation** |  |  |  |  |  |
| No | 8251 (56%) | 6663 (58%) | 5495 (57%) | 4189 (57%) | 4480 (56%) |
| Yes | 6258 (43%) | 4744 (41%) | 4032 (42%) | 3087 (42%) | 3502 (44%) |
| Unknown | 153 (1%) | 154 (1%) | 127 (1%) | 97 (1%) | 57 (1%) |
| **PSA Category, ng/mL** |  |  |  |  |  |
| <10 | 9328 (64%) | 7021 (61%) | 5747 (60%) | 4425 (60%) | 4682 (58%) |
| 10-20 | 1443 (10%) | 1150 (10%) | 1011 (10%) | 764 (10%) | 839 (10%) |
| ≥20 | 788 (5%) | 671 (6%) | 603 (6%) | 423 (6%) | 495 (6%) |
| Unknown | 3103 (21%) | 2719 (24%) | 2293 (24%) | 1761 (24%) | 2023 (25%) |
| **Neighborhood SES (Quintile)** |  |  |  |  |  |
| Quintile 1 (Most deprived) | 3114 (21%) | 1787 (15%) | 1237 (13%) | 998 (14%) | 1106 (14%) |
| Quintile 2 | 2843 (19%) | 2204 (19%) | 1614 (17%) | 1450 (20%) | 1413 (18%) |
| Quintile 3 | 2768 (19%) | 2403 (21%) | 1874 (19%) | 1409 (19%) | 1781 (22%) |
| Quintile 4 | 2871 (20%) | 2530 (22%) | 2534 (26%) | 1657 (22%) | 1830 (23%) |
| Quintile 5 (Least deprived) | 3066 (21%) | 2637 (23%) | 2395 (25%) | 1859 (25%) | 1909 (24%) |
| **County-level PSA, % (mean (SD))** | 0.57 (0.05) | 0.56 (0.05) | 0.56 (0.05) | 0.58 (0.03) | 0.59 (0.03) |
| Unknown | 1204 (8%) | 1547 (13%) | 1860 (19%) | 1860 (25%) | 2945 (37%) |
| **NO_2_, ppb (mean (SD))** | 25.85 (6.64) | 27.03 (6.24) | 27.84 (6.30) | 30.51 (7.67) | 32.54 (7.00) |
| **PM_2.5_, µg/m^3^ (mean (SD))** | 9.17 (1.37) | 9.24 (1.50) | 9.31 (1.75) | 10.08 (1.96) | 10.38 (2.20) |
| **NDVI 270m (mean (SD))** | 0.34 (0.17) | 0.34 (0.17) | 0.35 (0.17) | 0.33 (0.18) | 0.32 (0.17) |

Abbreviations: IQR=interquartile range; NDVI=Normalized Difference Vegetation Index; NO_2_=nitrogen dioxide; PFOS=perfluorooctane sulfonate, PM_2.5_=fine particulate matter ≤2.5 microns in diameter; PSA=prostate-specific antigen; SD=standard deviation; SES=socioeconomic status

**Supplementary Table 7. Characteristics of New Jersey Men with Prostate Cancer Stratified by Quintile of PFNA**

|  | **Categories of PFNA** | | |
| --- | --- | --- | --- |
| **Characteristic (n, %)** | **<2 ng/L** | **2-<3.8 ng/L** | **≥3.8 ng/L** |
| **N** | **50,542** | **372** | **375** |
| **PFNA, ng/L (median, [IQR])** | 1.01 (1.00, 1.02) | 3.31 (2.62, 3.61) | 6.39 (4.57, 9.50) |
| **Age at diagnosis, years (mean, SD)** | 67.00 (61.00, 74.00) | 66.00 (60.00, 72.00) | 66.00 (60.00, 74.00) |
| **Age at diagnosis, years** |  |  |  |
| <50 | 1714 (3%) | 12 (3%) | 17 (5%) |
| 50-60 | 10837 (21%) | 88 (24%) | 78 (21%) |
| 60-70 | 19831 (39%) | 155 (42%) | 143 (38%) |
| 70-80 | 13962 (28%) | 90 (24%) | 98 (26%) |
| ≥80 | 4198 (8%) | 27 (7%) | 39 (10%) |
| **Year of diagnosis** |  |  |  |
| 2006-2010 | 18405 (36%) | 163 (44%) | 134 (36%) |
| 2010-2014 | 16300 (32%) | 126 (34%) | 142 (38%) |
| 2014 -2017 | 15837 (31%) | 83 (22%) | 99 (26%) |
| **Race/Ethnicity** |  |  |  |
| NHW | 35481 (70%) | 165 (44%) | 257 (69%) |
| NHB | 8131 (16%) | 169 (45%) | 98 (26%) |
| NHAAPI | 1971 (4%) | 3 (1%) | 3 (1%) |
| Hispanic | 4959 (10%) | 35 (9%) | 17 (5%) |
| **Marital Status** |  |  |  |
| No | 10232 (20%) | 134 (36%) | 103 (27%) |
| Yes | 31690 (63%) | 197 (53%) | 225 (60%) |
| Missing | 8620 (17%) | 41 (11%) | 47 (13%) |
| **Insurance** |  |  |  |
| Uninsured | 3398 (9%) | 25 (8%) | 34 (11%) |
| Private | 14374 (38%) | 121 (38%) | 102 (32%) |
| Medicaid | 750 (2%) | 22 (7%) | 11 (4%) |
| Medicare | 18995 (50%) | 153 (48%) | 166 (53%) |
| Other government | 134 (0%) | 1 (0%) | 1 (0%) |
| Unknown | 12891 | 50 | 61 |
| **Gleason (combined)** |  |  |  |
| <7 | 27790 (60%) | 174 (52%) | 187 (55%) |
| ≥7 | 18567 (40%) | 160 (48%) | 156 (45%) |
| Unknown | 4185 | 38 | 32 |
| **Stage** |  |  |  |
| Localized | 34379 (87%) | 259 (84%) | 285 (88%) |
| Regional | 3288 (8%) | 29 (9%) | 25 (8%) |
| Distant | 1749 (4%) | 19 (6%) | 15 (5%) |
| Unknown | 11126 | 65 | 50 |
| **Surgery** |  |  |  |
| No | 33225 (67%) | 244 (68%) | 246 (66%) |
| Yes | 16372 (33%) | 117 (32%) | 124 (34%) |
| Unknown | 945 | 11 | 5 |
| **Hormone treatment** |  |  |  |
| No | 35739 (73%) | 234 (66%) | 255 (69%) |
| Yes | 13508 (27%) | 121 (34%) | 112 (31%) |
| Unknown | 1295 | 17 | 8 |
| **Radiation** |  |  |  |
| No | 28627 (57%) | 223 (61%) | 228 (61%) |
| Yes | 21336 (43%) | 142 (39%) | 145 (39%) |
| Unknown | 579 | 7 | 2 |
| **PSA Category, ng/mL** |  |  |  |
| <10 | 30760 (79%) | 213 (73%) | 230 (78%) |
| 10-20 | 5118 (13%) | 46 (16%) | 43 (15%) |
| ≥20 | 2928 (8%) | 31 (11%) | 21 (7%) |
| Unknown | 11736 | 82 | 81 |
| **Neighborhood SES (Quintile)** |  |  |  |
| Quintile 1 (Most deprived) | 7818 (15%) | 240 (65%) | 184 (49%) |
| Quintile 2 | 9346 (18%) | 57 (15%) | 121 (32%) |
| Quintile 3 | 10165 (20%) | 32 (9%) | 38 (10%) |
| Quintile 4 | 11371 (22%) | 38 (10%) | 13 (3%) |
| Quintile 5 (Least deprived) | 11842 (23%) | 5 (1%) | 19 (5%) |
| **County-level PSA, % (mean (SD))** | 0.58 (0.54, 0.60) | 0.60 (0.55, 0.60) | 0.59 (0.54, 0.60) |
| Unknown | 9338 (18%) | 46 (12%) | 32 (9%) |
| **NO_2_, ppb (mean (SD))** | 28.36 (23.40, 33.05) | 28.40 (24.44, 33.56) | 27.66 (23.57, 31.86) |
| **PM_2.5_, µg/m^3^ (mean (SD))** | 9.24 (8.27, 10.62) | 10.69 (9.80, 12.50) | 10.93 (9.73, 12.11) |
| **NDVI 270m (mean (SD))** | 0.35 (0.21, 0.48) | 0.31 (0.18, 0.43) | 0.32 (0.21, 0.44) |

Abbreviations: IQR=interquartile range; NDVI=Normalized Difference Vegetation Index; NO_2_=nitrogen dioxide; PFNA=perfluorononanoic acid, PM_2.5_=fine particulate matter ≤2.5 microns in diameter; PSA=prostate-specific antigen; SD=standard deviation; SES=socioeconomic status

**Supplementary Table 8. Associations of drinking water PFOA, PFOS and PFNA with prostate cancer mortality in New Jersey men with prostate cancer using inverse probability of censoring weights for competing risks**

|  |  | **Quintiles of Exposure** | | | | |  |
| --- | --- | --- | --- | --- | --- | --- | --- |
|  | **Continuous^a^** | **Q1** | **Q2** | **Q3** | **Q4** | **Q5** | ***P*_trend_** |
|  | **aHR (95% CI)** | **aHR (95% CI)** | **aHR (95% CI)** | **aHR (95% CI)** | **aHR (95% CI)** | **aHR (95% CI)** |  |
| PFOA |  |  |  |  |  |  |  |
| Cases/Person-years^b^ | 4,129/458,825 | 805/92,537 | 811/91,682 | 874/90,889 | 798/92010 | 841/91,707 |  |
| Minimal^c^ | 1.00 (0.97, 1.03) | Ref | 1.02 (0.92, 1.13) | 1.09 (0.98, 1.21) | 1.00 (0.90, 1.11) | 1.01 (0.91, 1.12) | 0.58 |
| Full^d^ | 1.01 (0.97, 1.04) | Ref | 0.97 (0.87, 1.08) | 1.07 (0.95, 1.19) | 0.95 (0.84, 1.06) | 0.97 (0.86, 1.10) | 0.49 |
| PFOS |  |  |  |  |  |  |  |
| Cases/Person-years^b^ | 4,129/458,935 | 835/92,706 | 831/92,124 | 769/91,891 | 825/91,092 | 870/91,122 |  |
| Minimal^c^ | 1.00 (0.98, 1.02) | Ref | 1.00 (0.91, 1.11) | 0.92 (0.83, 1.03) | 1.00 (0.90, 1.12) | 1.03 (0.92, 1.15) | 0.26 |
| Full^d^ | 1.00 (0.98, 1.02) | Ref | 0.99 (0.89, 1.11) | 0.95 (0.85, 1.06) | 0.95 (0.85, 1.07) | 1.01 (0.90, 1.15) | 0.49 |
| PFNA |  |  |  |  |  |  |  |
| Cases/Person-years^b^ | 4,128/458,695 | 4,073/455,082 | 28/1,915 | 27/1,789 | - | - |  |
| Minimal^c^ | 1.00 (1.00, 1.00) | Ref | 1.69 (1.15, 2.50) | 1.58 (1.04, 2.39) | - | - | 0.002 |
| Full^d^ | 1.00 (1.00, 1.00) | Ref | 1.17 (0.77, 1.78) | 1.37 (0.89, 2.11) | - | - | 0.10 |

^a^Per interquartile range, ^b^counts of cases and person-years estimated using inverse probability of censoring weights from first imputed dataset, ^c^Adjusted for age, diagnosis year, ^d^Adjusted for ^c^ and race/ethnicity, marital status, insurance status, stage, treatment (surgery and or radiation), PSA, nSES quintile, NO_2_, PM_2.5_, and NDVI (green space)

**Supplementary Table 9. Associations of drinking water PFOA, PFOS and PFNA with cardiovascular disease mortality in New Jersey men with prostate cancer using inverse probability of censoring weights for competing risks**

|  |  | **Quintiles of Exposure** | | | | |  |
| --- | --- | --- | --- | --- | --- | --- | --- |
|  | **Continuous^a^** | **Q1** | **Q2** | **Q3** | **Q4** | **Q5** | ***P*_trend_** |
|  | **aHR (95% CI)** | **aHR (95% CI)** | **aHR (95% CI)** | **aHR (95% CI)** | **aHR (95% CI)** | **aHR (95% CI)** |  |
| PFOA |  |  |  |  |  |  |  |
| Cases/Person-years | 5,544/455,095 | 980/91,831 | 1,121/91,019 | 1,164/90,041 | 1,128/90,951 | 1,152/90,951 |  |
| Minimal^b^ | 0.99 (0.97, 1.02) | Ref | 1.09 (0.99, 1.19) | 0.97 (0.88, 1.06) | 1.00 (0.91, 1.10) | 1.01 (0.92, 1.11) | 0.72 |
| Full^c^ | 1.02 (0.99, 1.04) | Ref | 1.10 (1.00, 1.21) | 1.02 (0.92, 1.12) | 1.05 (0.95, 1.15) | 1.12 (1.01, 1.24) | 0.09 |
| PFOS |  |  |  |  |  |  |  |
| Cases/Person-years | 5,547/455,259 | 927/92,077 | 982/91,500 | 1,107/91,033 | 1,294/90,316 | 1,236/90,333 |  |
| Minimal^b^ | 0.99 (0.97, 1.00) | Ref | 0.96 (0.87, 1.05) | 0.95 (0.86, 1.04) | 0.99 (0.90, 1.09) | 0.92 (0.83, 1.01) | 0.17 |
| Full^c^ | 1.01 (0.99, 1.03) | Ref | 1.00 (0.91, 1.10) | 1.00 (0.91, 1.10) | 1.04 (0.94, 1.15) | 1.03 (0.93, 1.15) | 0.49 |
| PFNA |  |  |  |  |  |  |  |
| Cases/Person-years | 5,540/454,918 | 5,500/451,307 | 21/1,902 | 20/1,772 | - | - |  |
| Minimal^b^ | 1.00 (1.00, 1.00) | Ref | 1.32 (0.86, 2.03) | 1.21 (0.77, 1.91) | - | - | 0.20 |
| Full^c^ | 1.00 (1.00, 1.00) | Ref | 1.06 (0.69, 1.64) | 1.00 (0.64, 1.58) | - | - | 0.90 |

^a^Per interquartile range, ^b^counts of cases and person-years estimated using inverse probability of censoring weights from first imputed dataset, ^c^Adjusted for age, diagnosis year, ^d^Adjusted for ^c^ and race/ethnicity, marital status, insurance status, stage, treatment (surgery and or radiation), PSA, nSES quintile, NO_2_, PM_2.5_, and NDVI (green space)

**Supplementary Table 10. Quantile g-computation results for mixtures of PFOA, PFOS and PFNA quintiles and all-cause, prostate cancer, and cardiovascular disease mortality in men with prostate cancer, New Jersey, 2006-2024.**

| **Per 1-quintile increase across PFOA, PFOS and PFNA** | **aHR (95% CI)** |
| --- | --- |
| All-cause mortality | 1.01 (0.99, 1.03) |
| Prostate cancer mortality | 1.02 (0.97, 1.07) |
| Cardiovascular mortality | 1.02 (0.98, 1.06) |

Adjusted for age, diagnosis year, race/ethnicity, marital status, insurance status, stage, treatment (surgery and or radiation), PSA, nSES quintile, NO_2_, PM_2.5_, and NDVI (green space)

**Supplementary Table 11. Associations of PFAS with mortality in prostate cancer survivors, stratified by race and ethnicity, age and state, New Jersey, 2006-2024.**

|  | **All-cause mortality** | | **Prostate cancer mortality** | | **Cardiovascular disease mortality** | |
| --- | --- | --- | --- | --- | --- | --- |
|  | **aHR (95% CI)** | ***P*_het_** | **aHR (95% CI)** | ***P*_het_** | **aHR (95% CI)** | ***P*_het_** |
| **PFOA** |  |  |  |  |  |  |
| **Race and ethnicity** |  | 0.73 |  | 0.49 |  | 0.59 |
| NHW | 1.00 (0.98, 1.02) |  | 1.00 (0.96, 1.04) |  | 1.02 (0.99, 1.06) |  |
| NHB | 0.99 (0.97, 1.02) |  | 1.04 (0.99, 1.08) |  | 1.00 (0.95, 1.05) |  |
| NHAAI | 0.98 (0.90, 1.06) |  | 1.02 (0.88, 1.18) |  | 0.93 (0.76, 1.13) |  |
| Hispanic | 1.02 (0.98, 1.06) |  | 0.97 (0.89, 1.06) |  | 1.00 (0.93, 1.09) |  |
| **Age** |  | 0.14 |  | 0.89 |  | 0.38 |
| <65 years | 0.98 (0.95, 1.01) |  | 1.01 (0.95, 1.06) |  | 0.99 (0.93, 1.06) |  |
| ≥65 years | 1.01 (0.99, 1.02) |  | 1.01 (0.98, 1.04) |  | 1.02 (1.00, 1.05) |  |
| **Stage at diagnosis** |  | 0.95 |  | 0.43 |  | 0.47 |
| Localized | 1.00 (0.99, 1.02) |  | 1.03 (0.99, 1.06) |  | 1.01 (0.98, 1.04) |  |
| Regional | 0.99 (0.94, 1.04) |  | 0.98 (0.90, 1.08) |  | 1.04 (0.93, 1.16) |  |
| Distant | 0.99 (0.95, 1.04) |  | 0.99 (0.94, 1.04) |  | 1.07 (0.98, 1.16) |  |
| **PFOS** |  |  |  |  |  |  |
| **Race and ethnicity** |  | 0.92 |  | 0.34 |  | 0.58 |
| NHW | 0.99 (0.98, 1.00) |  | 1.00 (0.97, 1.02) |  | 1.01 (0.99, 1.03) |  |
| NHB | 1.00 (0.98, 1.03) |  | 1.04 (0.99, 1.08) |  | 1.02 (0.97, 1.06) |  |
| NHAAI | 1.00 (0.95, 1.06) |  | 1.05 (0.96, 1.14) |  | 1.00 (0.88, 1.14) |  |
| Hispanic | 1.00 (0.97, 1.02) |  | 0.99 (0.93, 1.04) |  | 0.97 (0.91, 1.03) |  |
| **Age** |  | 0.81 |  | 0.30 |  | 0.92 |
| <65 years | 1.00 (0.98, 1.02) |  | 1.02 (0.98, 1.06) |  | 1.01 (0.96, 1.06) |  |
| ≥65 years | 1.00 (0.99, 1.01) |  | 1.00 (0.98, 1.02) |  | 1.01 (0.99, 1.03) |  |
| **Stage at diagnosis** |  | 0.84 |  | 0.12 |  | 0.55 |
| Localized | 1.00 (0.99, 1.01) |  | 1.03 (1.00, 1.05) |  | 1.00 (0.98, 1.02) |  |
| Regional | 0.99 (0.95, 1.03) |  | 1.01 (0.94, 1.07) |  | 0.99 (0.90, 1.09) |  |
| Distant | 0.98 (0.95, 1.01) |  | 0.97 (0.93, 1.01) |  | 1.04 (0.98, 1.11) |  |
| **PFNA** |  |  |  |  |  |  |
| **Race and ethnicity** |  | 0.26 |  | 1.00 |  | 0.40 |
| NHW | 1.00 (1.00, 1.00) |  | 1.00 (1.00, 1.00) |  | 1.00 (1.00, 1.00) |  |
| NHB | 1.00 (1.00, 1.00) |  | 1.00 (1.00, 1.00) |  | 1.00 (1.00, 1.00) |  |
| NHAAI | 1.00 (1.00, 1.00) |  | 1.00 (0.99, 1.01) |  | 0.99 (0.93, 1.05) |  |
| Hispanic | 1.00 (1.00, 1.00) |  | 1.00 (1.00, 1.00) |  | 1.00 (1.00, 1.00) |  |
| **Age** |  | 0.12 |  | 0.89 |  | 0.37 |
| <65 years | 1.00 (1.00, 1.00) |  | 1.00 (1.00, 1.00) |  | 1.00 (1.00, 1.00) |  |
| ≥65 years | 1.00 (1.00, 1.00) |  | 1.00 (1.00, 1.00) |  | 1.00 (1.00, 1.00) |  |
| **Stage at diagnosis** |  | 0.64 |  | 0.85 |  | 0.83 |
| Localized | 1.00 (1.00, 1.00) |  | 1.00 (1.00, 1.00) |  | 1.00 (1.00, 1.00) |  |
| Regional | 1.00 (1.00, 1.00) |  | 1.00 (1.00, 1.00) |  | 1.00 (1.00, 1.00) |  |
| Distant | 1.00 (1.00, 1.00) |  | 1.00 (1.00, 1.00) |  | 1.00 (1.00, 1.00) |  |

Adjusted for age, diagnosis year, race/ethnicity, marital status, insurance status, stage, treatment (surgery and or radiation), PSA, nSES quintile, NO_2_, PM_2.5_, and NDVI (green space)

**Supplementary Figure 1. Participant CONSORT Diagram**

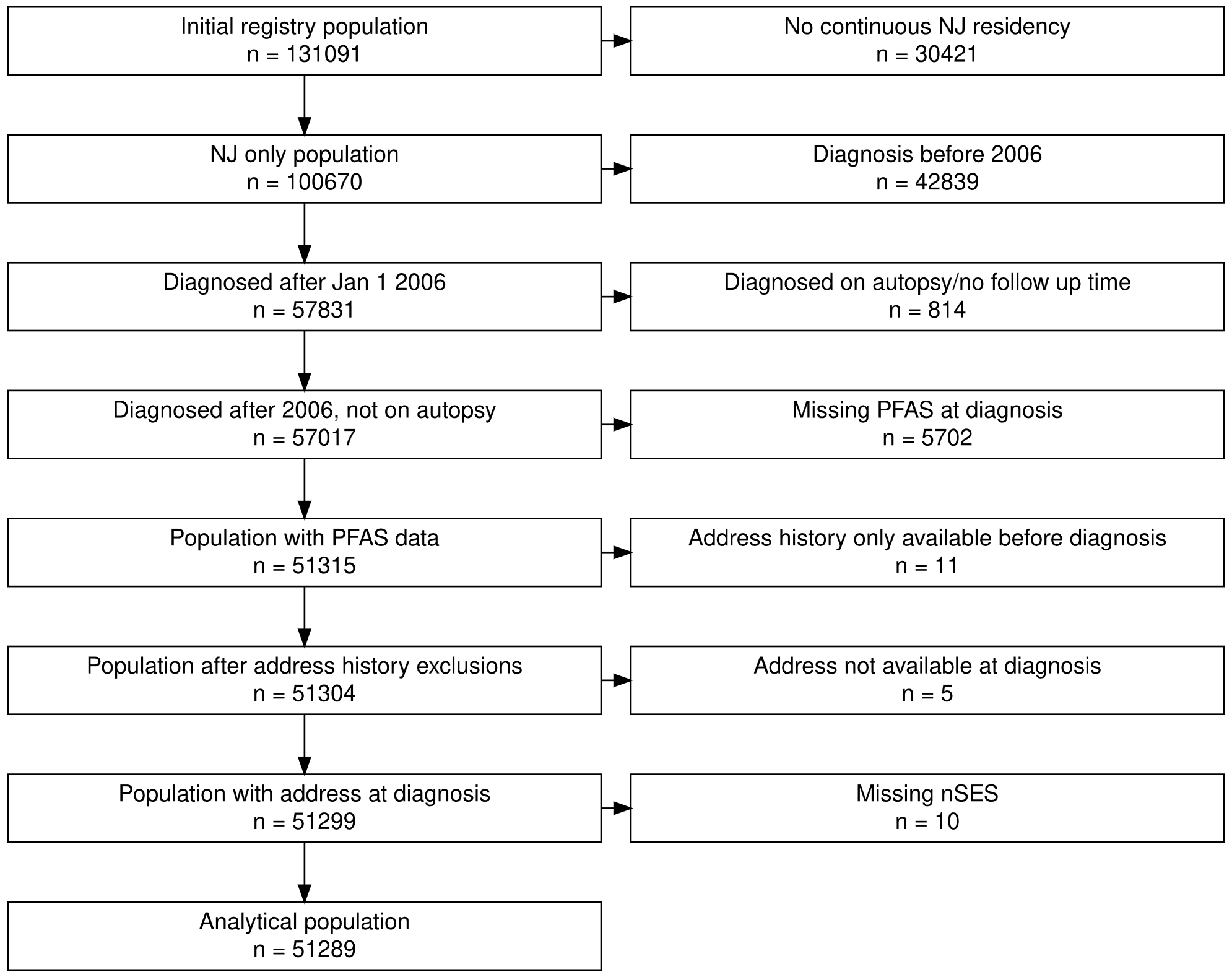

**Supplementary Figure 2. Correlation matrix for PFAS and neighborhood contextual factors and pollutants**
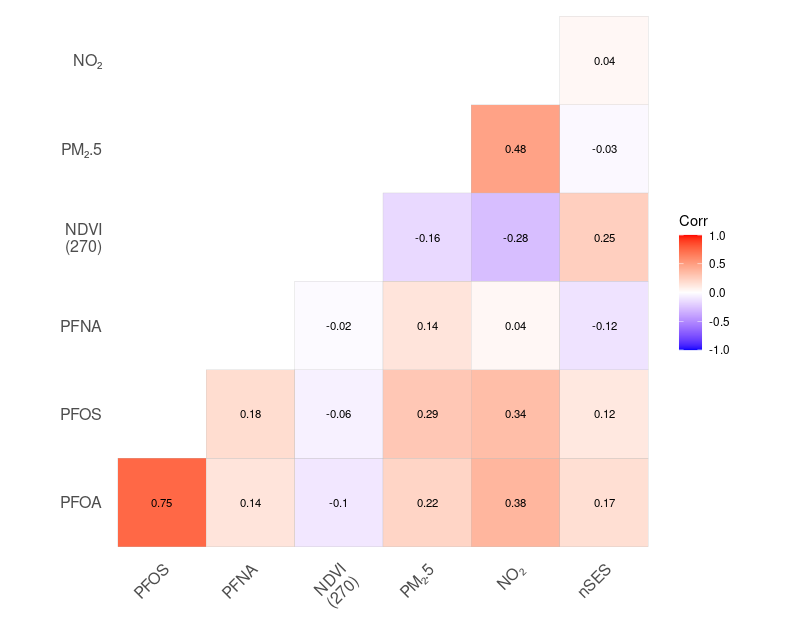

Abbreviations: PM_2.5_=fine particulate matter ≤2.5 microns in diameter, NO_2_=nitrogen dioxide, NDVI=Normalized Difference Vegetation Index within 270m, PFNA=perfluorononanoic acid, PFOS=perfluorooctane sulfonate, PFOA=perfluorooctanoic acid

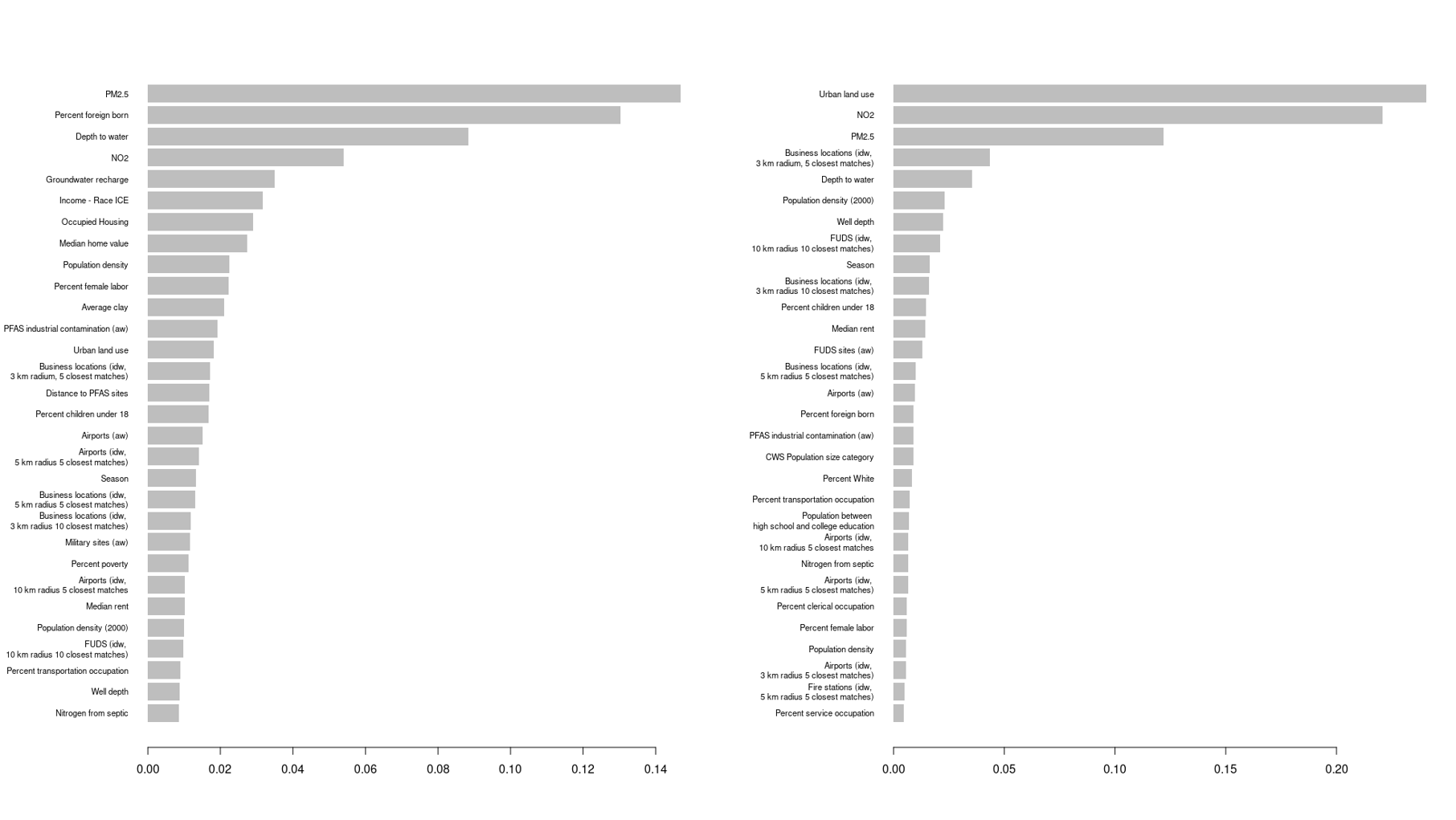
**Supplementary Figure 3. XGBoost Model for PFOA, Feature Importance (Top 30 predictors)**

**A**

**B**

Legend: A. Stage 1 (detect vs non-detect); B. Stage 2 (concentration). Abbreviations: ICE, Index of Concentrations at the Extremes; aw, average weight; idw, inverse distance weight; FUDS, Formerly Used Defense Sites Program; CWS, community water systems

B

A

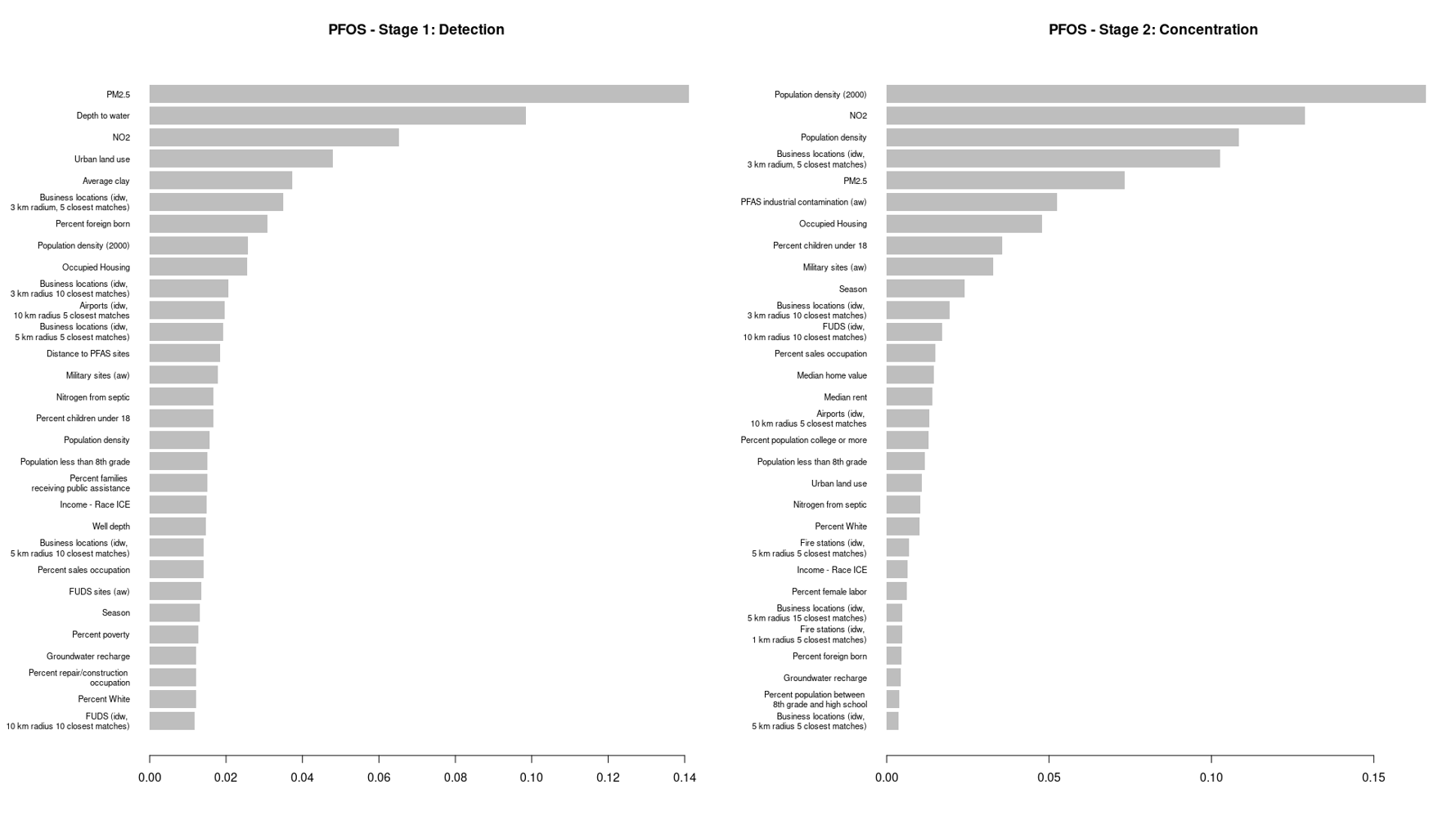
**Supplementary Figure 4. XGBoost Model for PFOS, Feature Importance (Top 30 predictors)**

**B**

**A**

Legend: A. Stage 1 (detect vs non-detect); B. Stage 2 (concentration). Abbreviations: ICE, Index of Concentrations at the Extremes; aw, average weight; idw, inverse distance weight; FUDS, Formerly Used Defense Sites Program; CWS, community water systems

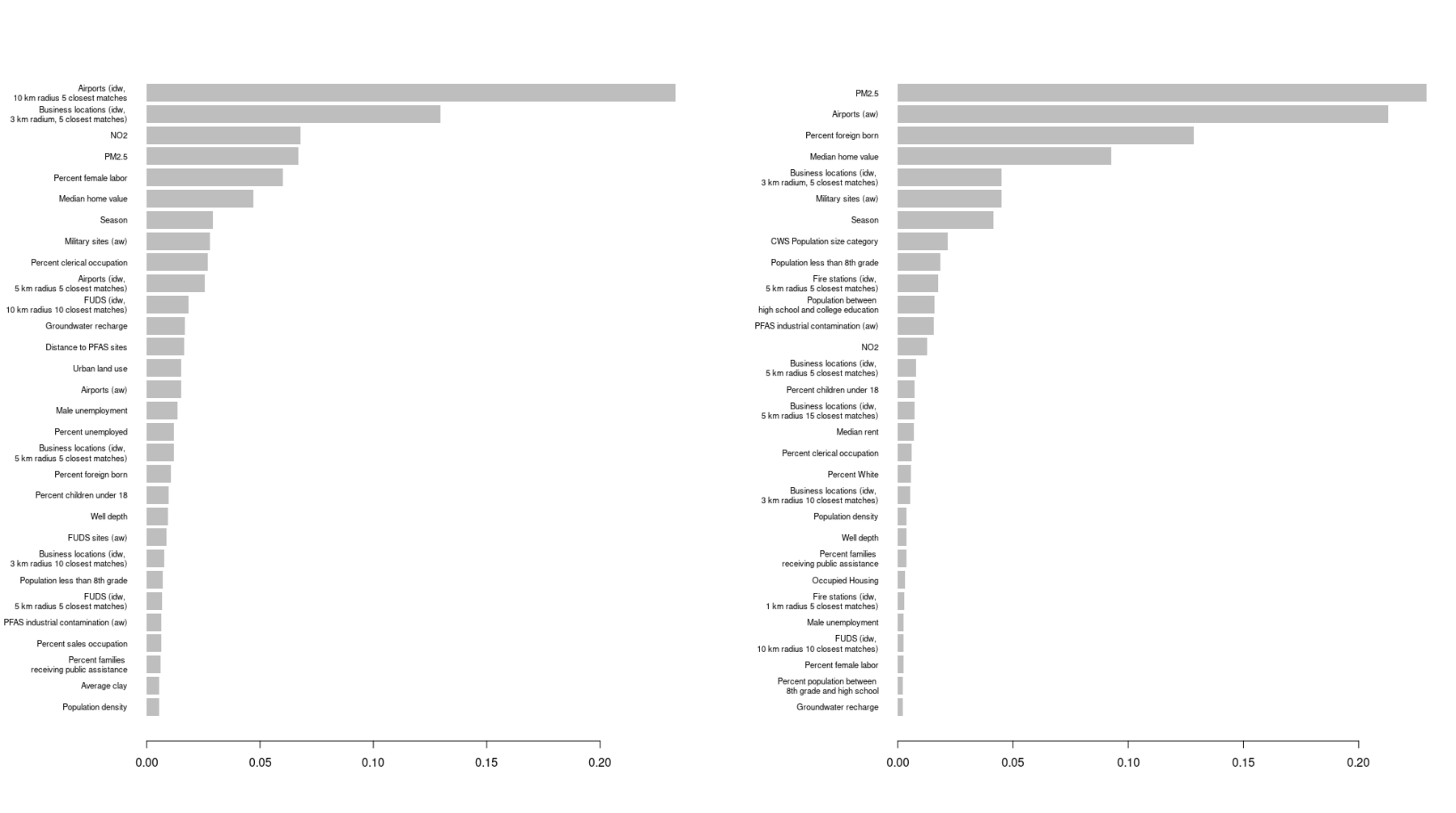
**Supplementary Figure 5. XBoost Model for PFNA, Feature Importance (Top 30 predictors)**

**B**

**A**

Legend: A. Stage 1 (detect vs non-detect); B. Stage 2 (concentration). Abbreviations: aw, average weight; idw, inverse distance weight; FUDS, Formerly Used Defense Sites Program; CWS, community water systems

**Supplementary Figure 6. Maps of predicted PFOA concentrations across 566 Community Water Systems, New Jersey, 2006-2025**

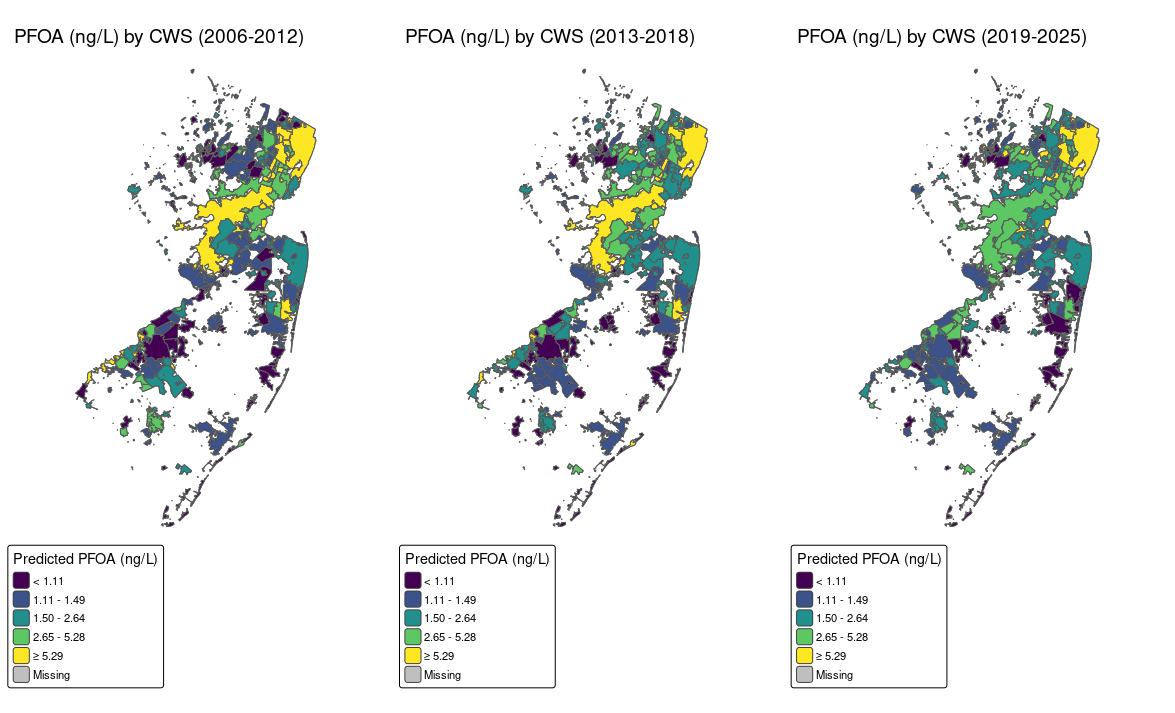

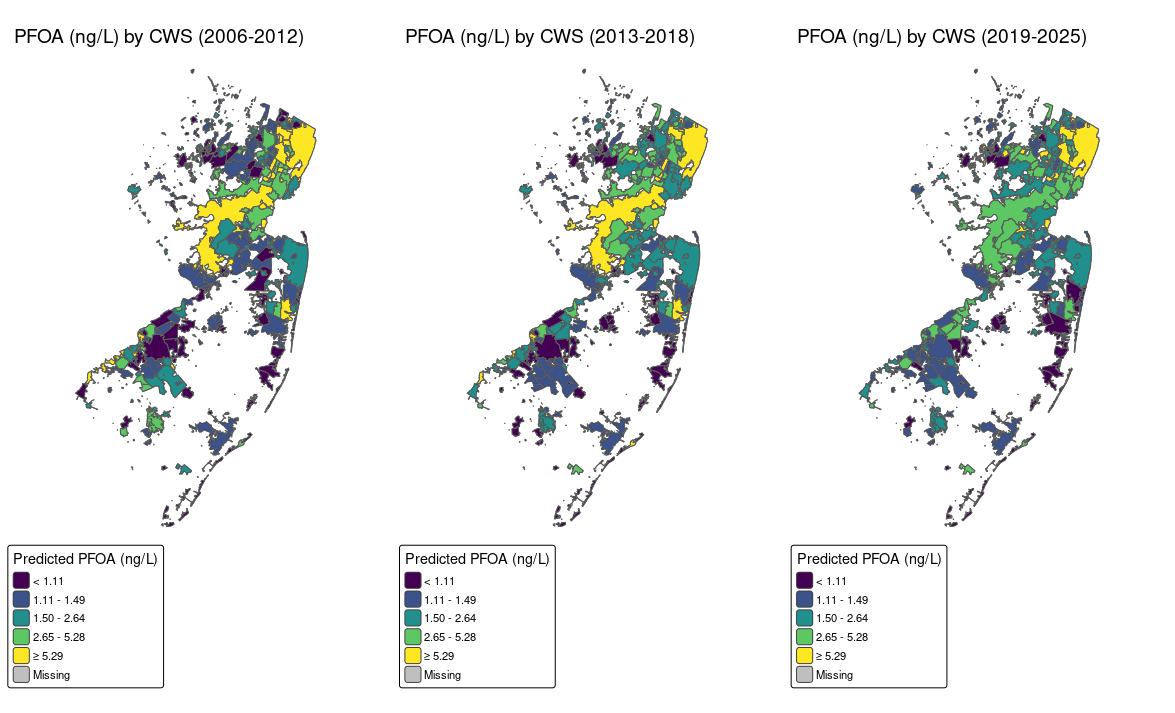

**Supplementary Figure 7. Maps of predicted PFOS concentrations across 566 Community Water Systems, New Jersey, 2006-2025**

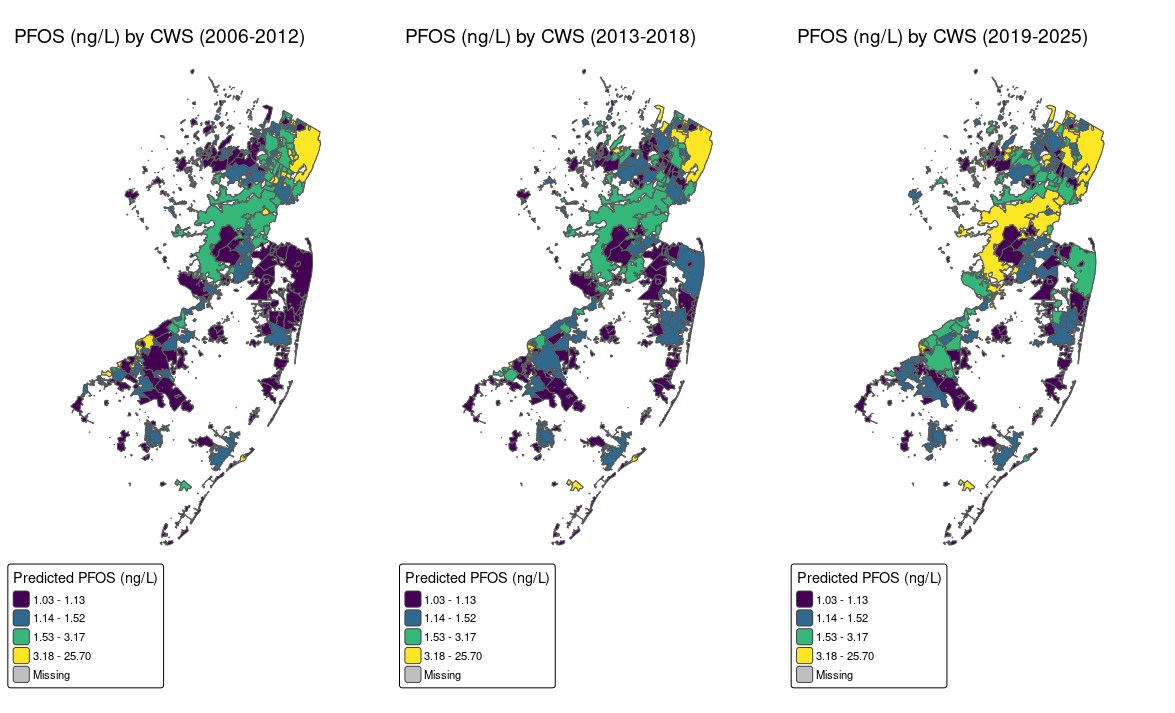

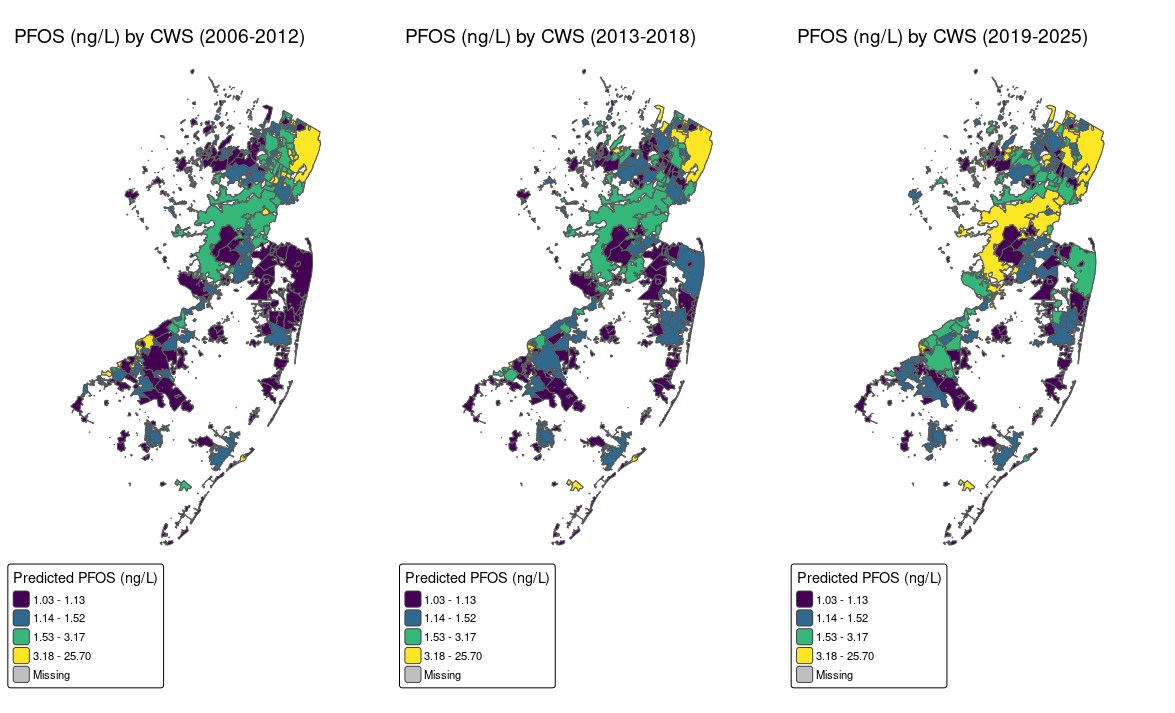

**Supplementary Figure 8. Maps of predicted PFNA concentrations across 566 Community Water Systems, New Jersey, 2006-2025**

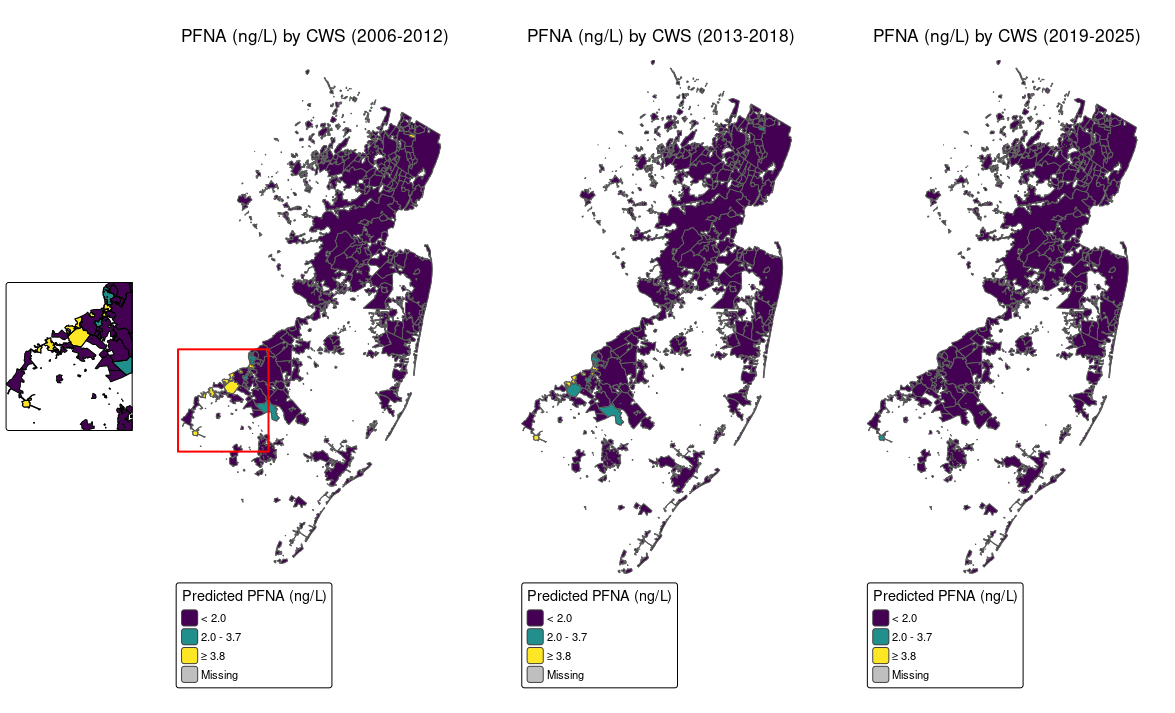

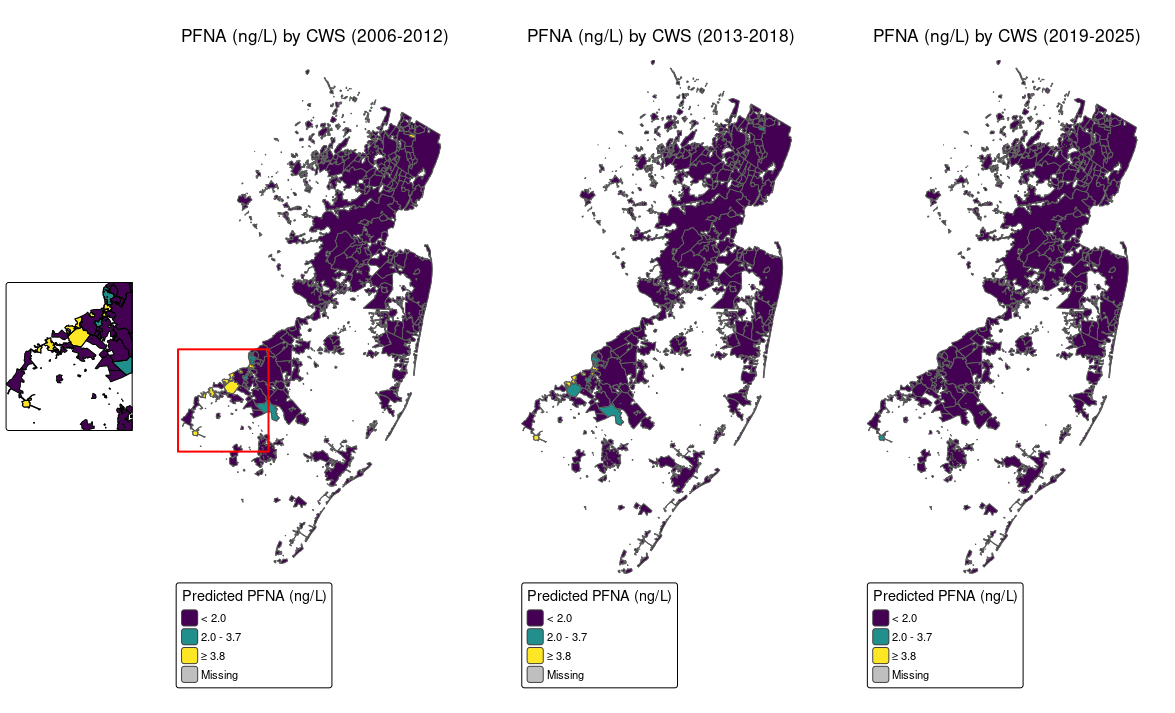

**Supplementary Figure 9. Weights for each PFAS from the quantile g-computation model**

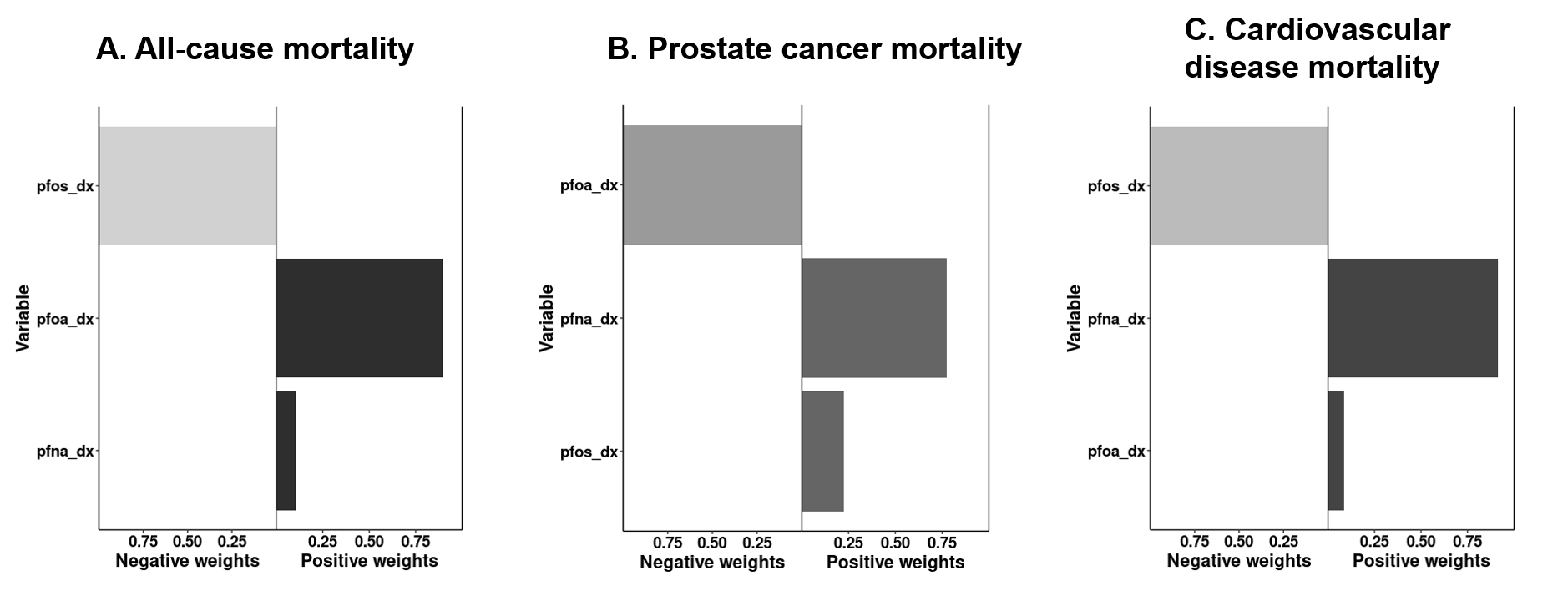

Legend: Abbreviations: pfoa_dx=PFOA, pfos_dx=PFOS, pfna_dx=PFNA. Weights represent the proportion of the positive or negative partial effect for each PFAS component in the mixture on all-cause (Panel A), prostate cancer (Panel B), and Cardiovascular Disease (Panel C) mortality. Shading indicates magnitude of effect size (darker is more positive).
